# Phthalates, Bisphenols, and Childhood Allergic Phenotypes: Findings from Two Birth Cohort Studies

**DOI:** 10.1101/2025.04.19.25325915

**Authors:** Thomas Boissiere-O’Neill, Nina Lazarevic, Anne-Louise Ponsonby, Peter D. Sly, Aimin Chen, Tamara L. Blake, Jeffrey R. Brook, Cassidy Du Berry, Louise King, Piushkumar J. Mandhane, Theo J. Moraes, Elinor Simons, Padmaja Subbarao, Dwan Vilcins, BIS, CHILD investigator groups

**Affiliations:** The University of Queensland, Child Health Research Centre, The Children’s Health and Environment Program QLD, 4101, Australia; National Centre for Epidemiology and Population Health, Australian National University, Canberra, ACT 2601, Australia; Florey Institute of Neuroscience and Mental Health, University of Melbourne, Melbourne, VIC 3010, Australia; Department of Biostatistics, Epidemiology and Informatics, Perelman School of Medicine, University of Pennsylvania, Philadelphia, PA 19104, United States; Dalla Lana School of Public Health, University of Toronto, Toronto, ON, Canada; Department of Respiratory and Sleep Medicine, Royal Children’s Hospital, Parkville, Victoria, Australia; Department of Paediatrics, University of Melbourne, Parkville, Victoria, Australia; Respiratory Group Murdoch Children’s Research Institute, Parkville, Victoria, Australia; Respiratory Research@Alfred, Monash University, Melbourne, Victoria, Australia; Department of Pediatrics, Faculty of Medicine and Dentistry, University of Alberta, Edmonton, Alberta, Canada; Department of Pediatrics & Translational Medicine, Hospital for Sick Children, University of Toronto, Toronto, Ontario, Canada; Department of Pediatrics and Child Health, Children’s Hospital Research Institute of Manitoba, University of Manitoba, Winnipeg, Manitoba, Canada; Department of Physiology, Faculty of Medicine, University of Toronto, Toronto, Canada

**Author notes:** **Correspondence**: Thomas Boissiere-O’Neill. BIS investigator group: Peter Vuillermin, Anne-Louise Ponsonby, Mimi L.K. Tang, Fiona Collier, Peter D. Sly, Leonard Harrison, Richard Saffery, Sarath Ranganathan, David Burgner, Toby Mansell and Martin O’Hely. CHILD investigator group: Padmaja Subbarao, Elinor Simons, Theo J. Moraes, Stuart E. Turvey, Piushkumar J. Mandhane and Meghan B. Azad.

**Keywords:** Phthalates, Bisphenols, Pregnancy, Childhood, Allergies, Atopy, Lung function

## Abstract

**Background:** Phthalates and bisphenols may contribute to childhood allergic outcomes, but whether these are differentially associated with atopic or non-atopic phenotypes is uncertain. We investigated whether early-life exposure to these chemicals differentially impacts atopic and non-atopic allergic outcomes.

**Methods:** We used two prospective birth cohorts to investigate distinct exposure windows. The Barwon Infant Study (n = 797) measured urinary phthalate and bisphenol metabolites at 36 weeks’ gestation. The Canadian Healthy Infant Longitudinal Development Study (n = 993) measured phthalate metabolites at 3, 12, and 36 months. Atopy was assessed via skin prick tests at 4-5 years. Outcomes included asthma, wheeze, eczema, and rhinitis at 4-5 years. Models were stratified by atopy. We modelled exposure mixtures using quantile G-computation and Bayesian kernel machine regression.

**Results:** Phthalate mixtures were associated with increased asthma risk in both exposure windows. Prenatal phthalate mixtures were more strongly associated with non-atopic asthma (adjusted risk ratio [aRR] = 1.83; 95% confidence interval [CI]: 1.10-3.04), with evidence of effect modification by atopy (p = 0.02 for interaction). Postnatal phthalate mixtures were associated with non-atopic asthma (aRR = 1.82, 95% CI: 1.19-2.78), though the association did not differ by phenotype (p = 0.45 for interaction). Phthalate mixtures showed U-shaped (prenatal) and inverse U-shaped (postnatal) associations with atopic asthma, and linear associations with non-atopic asthma. There was little evidence of associations for other allergic outcomes.

**Conclusion:** Early-life exposure to phthalates may differentially influence the risk of childhood atopic and non-atopic asthma. Future studies are needed to confirm these associations.

**HIGHLIGHTS:**

- We used two birth cohorts to examine distinct exposure windows to phthalates.
- Pre-and postnatal phthalate exposure was associated with increased asthma risk.
- Pre-and postnatal associations with asthma differed by atopic status.
- There was little evidence of association with other allergic outcomes.

## 1. INTRODUCTION

Childhood asthma and allergic diseases are heterogeneous conditions, comprising different phenotypes with distinct presentations and immunological profiles (1, 2). For instance, eczema can be classified as atopic dermatitis, characterised by immunoglobulin (IgE)-mediated hypersensitivity, or non-atopic dermatitis, which lacks systemic allergic sensitisation (2). While atopy contributes to the development of asthma and other allergic conditions, its attributable fraction rarely exceeds 50% in population-based studies (3), demonstrating the heterogeneity of these conditions and the importance of distinguishing phenotypes by atopic status.

Among environmental risk factors, phthalates and bisphenols have drawn increasing concern (4). These chemicals are ubiquitous in consumer products, such as food packaging, personal care products and thermal receipts, leading to widespread population exposure in high-income countries (5). Animal and in vitro studies suggest that phthalates and bisphenols may disrupt immune function by upregulating T-helper (Th) 2 cytokines, interacting with estrogenic pathways, and inducing oxidative stress, thereby offering biologically plausible pathways for their involvement in the development of allergic outcomes (4, 6).

Epidemiological evidence also suggests associations between phthalate and bisphenol exposure and childhood asthma and eczema, with systematic reviews reporting modest effect sizes (7–9). However, findings across studies remain heterogeneous, partly due to differences in exposure timing, outcome definitions, or population susceptibility. Importantly, few studies have examined whether these chemicals are differentially associated with atopic and non-atopic phenotypes of allergic conditions. One longitudinal birth cohort study reported that specific phthalates were associated with almost a twofold increased risk of eczema in atopic boys up to five years old but not in non-atopic boys (10). In a Korean cross-sectional study, serum IgE mediated approximately 15% of the association between mono-carboxynonyl phthalate and allergic rhinitis in children aged 3–17 years (11). Similarly, a Taiwanese cohort study reported that IgE levels mediated 70% of the association between bisphenol A and physician-diagnosed asthma in six-year-old children (12).

These studies suggest that atopic mechanisms may partially mediate the effects of plastic chemical exposures on allergic outcomes.

However, investigating these mechanisms in early childhood is challenging, in part due to difficulties in establishing accurate diagnoses. For instance, the heterogeneity of preschool asthma often leads to diagnostic delays and uncertainty (13). Thus, lung function assessments offer an objective means to identify and quantify early or subclinical respiratory impairment. Yet, most epidemiological studies on phthalates, bisphenols, and lung function have focused on school-aged children or older populations (14), leaving a gap in the understanding of early-life respiratory effects of these chemicals. In preschool-aged children, tidal breathing techniques, such as multiple-breath washout (MBW) and oscillometry, offer a more feasible (15), and sensitive alternative to effort-based techniques, like spirometry, can be difficult to perform in pre-school aged children (16, 17)

Overall, important gaps remain. First, the extent to which associations with asthma and allergic outcomes differ by atopic status is understudied. Second, only few studies have evaluated both prenatal and postnatal exposures to phthalates. Third, limited work has applied lung function techniques in early pre-school children to investigate subclinical respiratory effects of these exposures.

In this study, we aimed to investigate whether early-life exposure to phthalates and bisphenols is differentially associated with atopic and non-atopic phenotypes of asthma and allergic outcomes in preschool-aged children. To address this, we leveraged two prospective birth cohorts: one in Australia, assessing prenatal exposures, and one in Canada, assessing postnatal exposures. We further investigated the associations between these chemicals and food allergy, as well as lung function, evaluated via ventilation inhomogeneity, resistance and reactance.

## 2. METHODS

### 2.1. Study population

In this study, two birth cohorts were used to examine distinct exposure windows: the Barwon Infant Study (BIS) to assess prenatal exposure to phthalates and bisphenols, and the Canadian Healthy Infant Longitudinal Development (CHILD) cohort to assess postnatal exposure to phthalates. Detailed descriptions of inclusion and exclusion criteria have been previously published (18, 19). The Barwon Infant Study is a population-based longitudinal cohort study with 1,074 mother-infant pairs recruited from the Barwon region in Victoria, Australia (18). Pregnant women at up to 28 weeks’ gestation were recruited if they were above 18 years, while children born <32 weeks’ gestation were excluded from the study. Ethics approval was granted by the Barwon Health Human Research and Ethics (HREC 10/24), and parents or legal guardians provided informed consent (18). The CHILD study included 3,542 infants born after 35 weeks’ gestation, with pregnant mothers recruited during the 2^nd^ or 3^rd^ trimester in four Canadian communities (Vancouver, Edmonton, Winnipeg, Toronto). The Human Research Ethics Boards at the Hospital for Sick Children, McMaster University, and the Universities of British Columbia, Manitoba, and Alberta approved the study, and parents or legal guardians provided informed consent (19). In both cohorts, we excluded children with missing exposure and outcome data (Figure S1). In the CHILD cohort, we also excluded children with only one urine sample collected (N = 503) to mitigate measurement error of the exposures. The methodology used in this study is shown in Figure 1 and described below.

**Figure 1.**
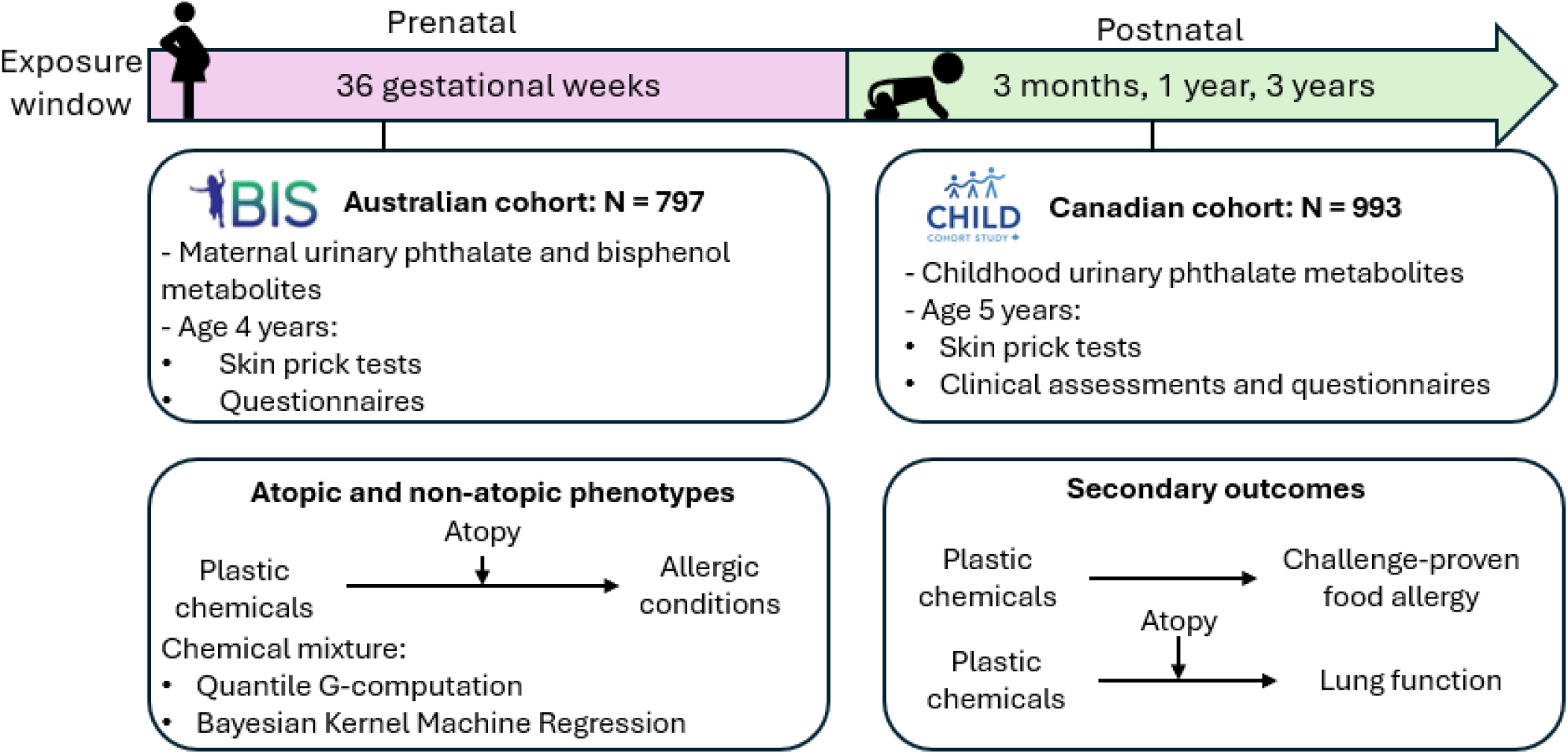
Overview of the methodology used in this study

### 2.2. Exposure

#### 2.2.1. Urinary metabolites measurements

Exposure was assessed using urinary metabolite concentrations in both cohorts, with prenatal exposure measured in BIS and postnatal exposure measured in CHILD. To ensure methodological comparability with previous studies from BIS and CHILD, we used the same methods for imputing values below the limit of detection (LOD) as used in those studies (20, 21).

In BIS, urine samples were collected from 842 pregnant women at approximately 36 weeks of gestation (mean = 36.0 weeks, SD = 0.7 weeks), on average three weeks before delivery (20). Samples were collected in glass or polypropylene containers, aliquoted, and stored at-80°C before shipment to the Queensland Alliance for Environmental Health Sciences for instrumental analysis (22). In CHILD, 1,578 children provided at least one urine sample at 3, 12, or 36 months of age. For non-toilet-trained children, urine was collected using absorbent pads and plastic film, then extracted and transferred to cryovials for storage at –80°C (21). Samples were analysed at AXYS Analytical Services Inc. using the Kato method (23).

Metabolites included in BIS were mono-methyl phthalate (MMP), mono-ethyl phthalate (MEP), mono-isobutyl phthalate (MiBP), mono-n-butyl phthalate (MnBP), mono-benzyl phthalate (MBzP), mono(2-ethyl-5-oxohexyl) phthalate (MEOHP), mono(2-ethyl-5-hydroxyhexyl) phthalate (MEHHP), mono(5-carboxy-2-ethylpentyl) phthalate (MECPP), mono(3-carboxpropyl) phthalate (MCPP), bisphenol A (BPA), bisphenol S (BPS) and bisphenol F (BPF). In CHILD, the metabolites included were MEP, MiBP, MnBP, MBzP, MEHP, MEOHP, MEHHP, and MCPP.

In BIS, phthalates with values under the LOD were imputed with LOD/√2 (20), while in CHILD, robust regression on order statistics was used (21). Due to bisphenols’ higher proportion of values below LOD, BPA was dichotomised, using the 75^th^ percentile as a cut-off, and the 90th percentile for BPS and BPF (Table S1), consistent with prior analyses in this cohort aimed at minimising bias from non-detected values and accounting for potential variability across analytical methods (24). In both cohorts, metabolites were corrected for specific gravity, using the Levine-Fahey equation. In BIS, phthalate metabolites were further standardised for batch effect and time of day at urine collection (20); this information was not available in CHILD (21). Metabolite distributions and detection frequencies are shown in Table S1.

#### 2.2.2. Daily intake estimation

Phthalate diester estimated daily intake was calculated to standardise metabolites for body weight, considering urinary fractional excretion, molecular-weight ratio, average daily urinary output and weight at sampling (20, 25):

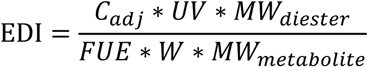

Here, *C_adj_* represents the standardised metabolite concentration (µg/L), *UV* denotes the average daily urine output (1.6 L during pregnancy)(20), *FUE* refers to the fraction of the metabolite excreted in urine, *MW* is the molecular weight (g/mol), and *W* corresponds to the individual’s body weight at the time of sampling (kg). In CHILD, UV/W was estimated from the child’s age at sampling (25). Urinary MMP was used to estimate the daily intake of dimethyl phthalate (DMP), MEP for diethyl phthalate (DEP), MnBP for di-n-butyl phthalate (DnBP), MiBP for di-isobutyl phthalate (DiBP), MBzP for butyl benzyl phthalate (BBzP), and MEHP, MEHHP, MEOHP, and MECPP for di(2-ethylhexyl) phthalate (DEHP). DiBP and DnBP were summed to derive the total dibutyl phthalate exposure (DBP). In CHILD, the daily intake for each phthalate was estimated at 3, 12 and 36 months, and the per-child average was used to represent exposure from infancy through early childhood. Since MCPP is a non-specific metabolite of high-molecular-weight phthalates (26), we did not calculate its daily intake. In BIS, measurements at 36 weeks’ gestation are referred to as *prenatal exposure*, whereas in CHILD, the average across time points is referred to as *postnatal exposure*.

### 2.3. Outcomes

In BIS, all outcomes were assessed at the age of four, whereas in CHILD, assessments were conducted at the age of five.

#### 2.3.1. Primary outcomes

Skin prick testing was employed to determine atopic status, defined as the presence of a wheal at least 3 mm larger than the negative control for any tested allergen, in the presence of a positive control ≥ 3 mm and a negative control ≤ 3 mm (27). The list of allergens tested in each cohort is provided in Table S2. In BIS, asthma was identified through parental report (“*Has your child ever had asthma?*”), and wheeze was assessed with the question, “*Has your child had wheezing or whistling from the chest in the last 12 months?*”. Eczema was defined using the modified UK Working Party criteria (28), and rhinitis was assessed through the question, “*Has your child ever had hay fever?*”. In CHILD, asthma, eczema, and rhinitis were assessed by subspecialist paediatricians and classified as “definite”, “possible,” or “no” (29). Only children with a “definite” diagnosis were considered cases. Wheeze was defined based on a positive response to the question, “*In the last year, has your child had a wheezing noise (whistling sound) coming from his/her chest either WITH a cold or WITHOUT a cold?*”.

#### 2.3.2. Secondary outcomes

To further investigate IgE-mediated outcomes, we investigated the association between prenatal exposures and food allergy in the BIS cohort. Food allergy was assessed at the one-year follow-up using a hospital-based open food challenge (27). Challenge-proven food allergy data was not available in CHILD. To assess early-life lung function, we evaluated ventilation inhomogeneity and lung volumes using multiple-breath washout (MBW) and reactance and resistance using oscillometry. MBW is a non-invasive tidal-breathing technique that measures ventilation distribution by quantifying the efficiency with which an inert gas is cleared from the lungs (30, 31). The lung clearance index (LCI_2.5_) indicates the number of lung volume turnovers necessary to expel the inert gas. Functional residual capacity (FRC), the volume of air remaining in the lungs after a normal expiration, is also derived from this test. Oscillometry, in contrast, applies external pressure oscillations at multiple frequencies during tidal breathing to assess airway mechanics (32). It quantifies respiratory impedance, which consists of resistance (R) and reactance (X). Resistance reflects energy loss due to airflow obstruction, while reactance captures the elastic and inertial properties of the respiratory system.

In both cohorts, MBW was performed using 2.5% nitrogen (N_2_) and an ultrasonic flowmeter (Exhalyzer D, Ecomedics, Duernten, Switzerland), with data analysed in Spiroware 3.3.1, incorporating cross-sensitivity corrections for the oxygen and carbon dioxide sensors (33). Testing was performed and analysed following current guidelines (30, 31). In BIS, lung function was also assessed with intrabreath oscillometry using equipment provided by the INCIRCLE collaboration (34). Oscillatory signals at 10 Hz were applied every 0.1 seconds during undisturbed tidal breathing, with impedance measured at end-inspiration and end-expiration (35). In CHILD, spectral oscillometry was performed using the TremoFlo system (THORASYS Thoracic Medical Systems Inc., Montreal, Canada), which delivered oscillations across a frequency range of 5 to 37 Hz (19). Due to differences in oscillometry techniques and to improve comparability between cohorts, resistance and reactance values in BIS were calculated as the mean of end-inspiratory and end-expiratory values at 10 Hz (R10 and X10), while in CHILD, values at 11 Hz (R11 and X11) were used.

### 2.4. Sociodemographic and risk factors

We first selected variables using a Directed Acyclic Graph (Figure S2), informed by prior knowledge and previous studies (4, 7). Definitions and cohort availability of each variable are shown in Table S3. Sociodemographic factors included maternal age at conception, marital status, remoteness area, caregiver education, household income, and child’s ethnicity. The Socio-Economic Indexes for Areas (SEIFA) Index of Relative Socioeconomic Disadvantage, a composite measure of area-based socioeconomic status, was also included as tertiles in BIS (36). Perinatal and birth characteristics included year and season of birth, birthweight, sex, and breastfeeding duration. Family and environmental factors included family history of asthma, tobacco smoke exposure during pregnancy, smoking inside the house during childhood, home renovation, household size, and pet ownership. In BIS, maternal diet during pregnancy was assessed using a food frequency questionnaire and principal component analysis (20, 37). This analysis identified three dietary patterns: (i) a modern healthy diet, (ii) a western unhealthy diet, and (iii) a traditional Anglo-Saxon diet. Factor scores were categorised into tertiles (low, medium, high), reflecting diet adherence. We also incorporated a volatile household product use score in BIS, based on the frequency of household product use during pregnancy, as previously developed by the Avon Longitudinal Study of Parents And Children (ALSPAC) cohort (20, 38).

### 2.5. Statistical analysis

Population characteristics and exposure concentrations were summarised using descriptive statistics. Since exposure variables followed a log-normal distribution, we applied a log2 transformation. Missing covariate data were imputed using multiple imputation by chained equations, with five datasets and 20 iterations (39). Given differences in exposure measurements and outcome assessments between BIS and CHILD, estimates were not pooled between cohorts.

#### 2.5.1. Atopic and non-atopic phenotypes

To evaluate potential nonlinearity, we first fitted generalised linear models with and without restricted cubic spline models (three knots at the 10th, 50th, and 90th percentiles), selecting the best-fit model using the Akaike Information Criterion (AIC) (40). Prenatal DEP in BIS, showed evidence of nonlinearity across several outcomes (Figure S3). We followed a systematic approach to build the multivariable models (41). First, we screened variables identified in the DAG as potential confounders. Second, variables that altered the exposure-outcome estimate by 10% or more in bivariable models were retained (41). Variables on the causal pathway (i.e., mediators) were excluded from adjustment. Associations between each exposure and binary outcomes were estimated using modified Poisson regression with robust standard errors to estimate crude and adjusted risk ratios (RR) with 95% confidence intervals (95% CI) (42). Continuous outcomes were scaled to enable between-model comparison, and associations were estimated with linear regression. We considered atopy and allergic outcomes separately and conducted stratified analyses by atopy, as previously recommended (43). Thus, we introduced interaction terms between each exposure and atopy to formally test for effect measure modification and derived phenotype-specific estimates, using the *marginaleffects* package (44). To estimate the combined effect of exposures, we applied quantile g-computation, showing RR and 95% CI associated with a one-quartile increase in all exposures (45). For consistency, the mixture analysis included only chemicals measured in both cohorts, excluding DMP and bisphenols from BIS. Interaction terms between the exposure mixture and atopy were added using the qgcompint package (46). Weights were averaged across imputed datasets to obtain pooled weights given the use of multiple imputation.

#### 2.5.2. Supplementary analyses

Outcome antecedents that did not substantially alter exposure-outcome associations were added to assess potential residual confounding. To confirm the results of quantile G-computation, we ran Bayesian Kernel Machine Regression (BKMR) on primary outcomes, for 50,000 iterations, stratified by atopic status. This method employs a Gaussian kernel function to estimate exposure effects by measuring distances between individuals in the exposure space. Prior distributions for the smoothing parameter (r) were identified using frequentist kernel machine regression, and uninformative priors were used for the remaining parameters in the final model (47). We adjusted the Metropolis-Hastings parameters (r_jump_, λ_jump_) to achieve acceptance rates of 20-40%. BKMR models were fitted within each imputed dataset and pooled using Rubin’s rule. Because skin prick test results can be influenced by factors such as pet ownership and allergen avoidance, we conducted a sensitivity analysis focusing on house dust mite and grass pollen from skin prick testing (Table S2), as these allergens are ubiquitous and less affected by modifiable factors (48, 49). To further avoid confounding by concomitant sensitisation, we used mutually exclusive categories: no sensitisation to HDM or grass (reference), sensitisation to HDM only, sensitisation to grass only, and sensitisation to both HDM and grass (50).

## 3. RESULTS

### 3.1. Sample characteristics

A total of 797 children from BIS and 993 from CHILD were included (Figure S1). There were no substantial differences in outcome and exposure distributions between study samples and full cohorts (Table S4-S5). Out of 467 children in BIS who underwent skin prick testing, 28.7% (n = 134) were atopic. In CHILD, out of 937 children, 13.1% (n = 123) were atopic. In both cohorts, atopic children were more likely to be males, to have a family history of asthma, and had higher outcome prevalence compared to non-atopic children (Table 1-2).) BIS had a lower proportion of university-educated caregivers, a lower rate of tobacco smoke exposure during pregnancy, smaller households, and a higher proportion of Caucasian/White children compared to CHILD. Overall, allergic outcomes were more prevalent in BIS compared to CHILD, although the proportion of children with eczema was similar between cohorts.

**Table 1.**
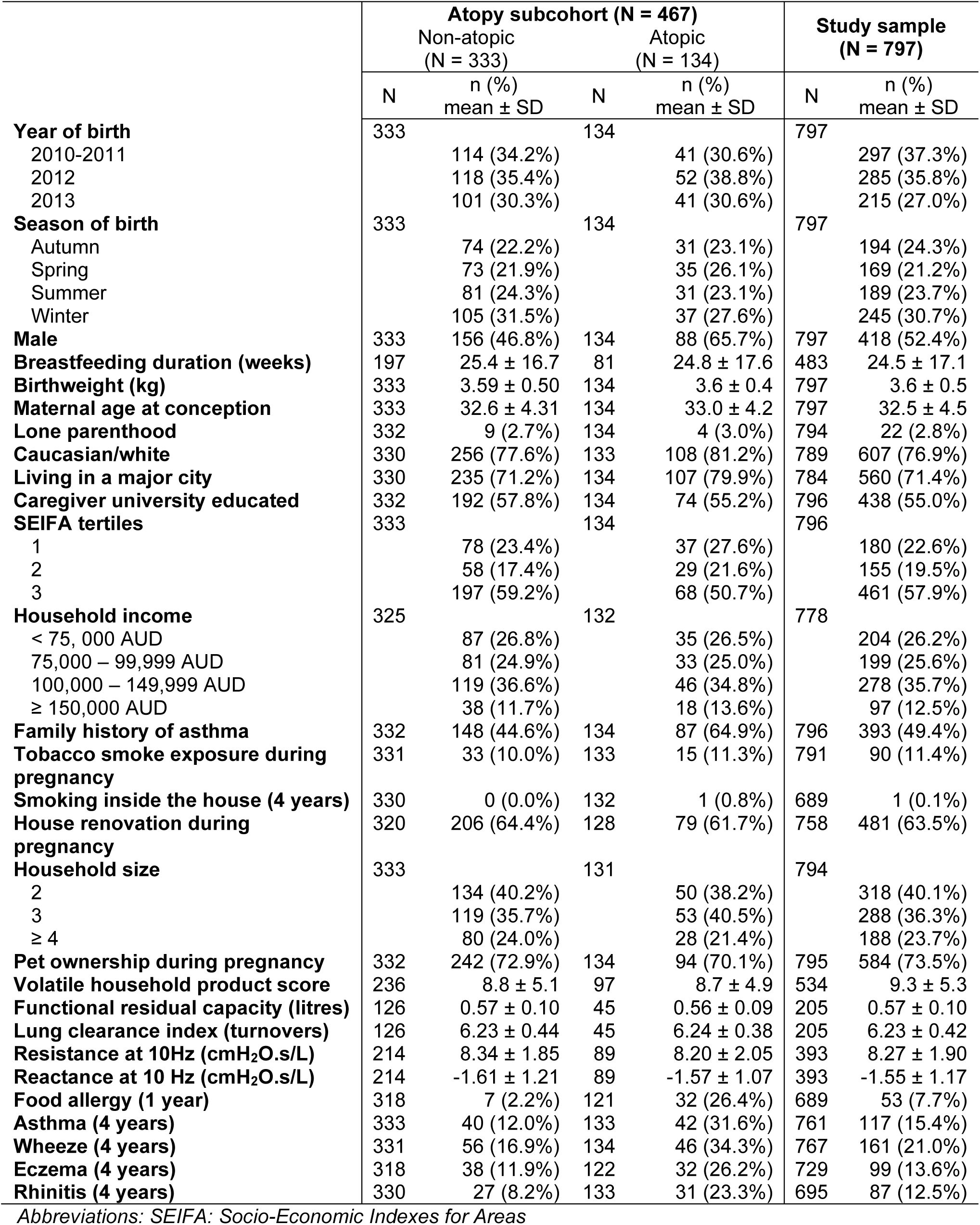
Descriptive characteristics of the Barwon Infant Study.

|  | Atopy subcohort (N = 467) |  |  |  | Study sample (N = 797) |  |
| --- | --- | --- | --- | --- | --- | --- |
|  | Non-atopic (N = 333) |  | Atopic (N = 134) |  |  |  |
|  | N | n (%)<br>mean ± SD | N | n (%)<br>mean ± SD | N | n (%)<br>mean ± SD |
| **Year of birth** | 333 |  | 134 |  | 797 |  |
| 2010-2011 |  | 114 (34.2%) |  | 41 (30.6%) |  | 297 (37.3%) |
| 2012 |  | 118 (35.4%) |  | 52 (38.8%) |  | 285 (35.8%) |
| 2013 |  | 101 (30.3%) |  | 41 (30.6%) |  | 215 (27.0%) |
| **Season of birth** | 333 |  | 134 |  | 797 |  |
| Autumn |  | 74 (22.2%) |  | 31 (23.1%) |  | 194 (24.3%) |
| Spring |  | 73 (21.9%) |  | 35 (26.1%) |  | 169 (21.2%) |
| Summer |  | 81 (24.3%) |  | 31 (23.1%) |  | 189 (23.7%) |
| Winter |  | 105 (31.5%) |  | 37 (27.6%) |  | 245 (30.7%) |
| **Male** | 333 | 156 (46.8%) | 134 | 88 (65.7%) | 797 | 418 (52.4%) |
| **Breastfeeding duration (weeks)** | 197 | 25.4 ± 16.7 | 81 | 24.8 ± 17.6 | 483 | 24.5 ± 17.1 |
| **Birthweight (kg)** | 333 | 3.59 ± 0.50 | 134 | 3.6 ± 0.4 | 797 | 3.6 ± 0.5 |
| **Maternal age at conception** | 333 | 32.6 ± 4.31 | 134 | 33.0 ± 4.2 | 797 | 32.5 ± 4.5 |
| **Lone parenthood** | 332 | 9 (2.7%) | 134 | 4 (3.0%) | 794 | 22 (2.8%) |
| **Caucasian/white** | 330 | 256 (77.6%) | 133 | 108 (81.2%) | 789 | 607 (76.9%) |
| **Living in a major city** | 330 | 235 (71.2%) | 134 | 107 (79.9%) | 784 | 560 (71.4%) |
| **Caregiver university educated** | 332 | 192 (57.8%) | 134 | 74 (55.2%) | 796 | 438 (55.0%) |
| **SEIFA tertiles** | 333 |  | 134 |  | 796 |  |
| 1 |  | 78 (23.4%) |  | 37 (27.6%) |  | 180 (22.6%) |
| 2 |  | 58 (17.4%) |  | 29 (21.6%) |  | 155 (19.5%) |
| 3 |  | 197 (59.2%) |  | 68 (50.7%) |  | 461 (57.9%) |
| **Household income** | 325 |  | 132 |  | 778 |  |
| < 75,000 AUD |  | 87 (26.8%) |  | 35 (26.5%) |  | 204 (26.2%) |
| 75,000 – 99,999 AUD |  | 81 (24.9%) |  | 33 (25.0%) |  | 199 (25.6%) |
| 100,000 – 149,999 AUD |  | 119 (36.6%) |  | 46 (34.8%) |  | 278 (35.7%) |
| ≥ 150,000 AUD |  | 38 (11.7%) |  | 18 (13.6%) |  | 97 (12.5%) |
| **Family history of asthma** | 332 | 148 (44.6%) | 134 | 87 (64.9%) | 796 | 393 (49.4%) |
| **Tobacco smoke exposure during pregnancy** | 331 | 33 (10.0%) | 133 | 15 (11.3%) | 791 | 90 (11.4%) |
| **Smoking inside the house (4 years)** | 330 | 0 (0.0%) | 132 | 1 (0.8%) | 689 | 1 (0.1%) |
| **House renovation during pregnancy** | 320 | 206 (64.4%) | 128 | 79 (61.7%) | 758 | 481 (63.5%) |
| **Household size** | 333 |  | 131 |  | 794 |  |
| 2 |  | 134 (40.2%) |  | 50 (38.2%) |  | 318 (40.1%) |
| 3 |  | 119 (35.7%) |  | 53 (40.5%) |  | 288 (36.3%) |
| ≥ 4 |  | 80 (24.0%) |  | 28 (21.4%) |  | 188 (23.7%) |
| **Pet ownership during pregnancy** | 332 | 242 (72.9%) | 134 | 94 (70.1%) | 795 | 584 (73.5%) |
| **Volatile household product score** | 236 | 8.8 ± 5.1 | 97 | 8.7 ± 4.9 | 534 | 9.3 ± 5.3 |
| **Functional residual capacity (litres)** | 126 | 0.57 ± 0.10 | 45 | 0.56 ± 0.09 | 205 | 0.57 ± 0.10 |
| **Lung clearance index (turnovers)** | 126 | 6.23 ± 0.44 | 45 | 6.24 ± 0.38 | 205 | 6.23 ± 0.42 |
| **Resistance at 10Hz (cmH<sub>2</sub>O.s/L)** | 214 | 8.34 ± 1.85 | 89 | 8.20 ± 2.05 | 393 | 8.27 ± 1.90 |
| **Reactance at 10 Hz (cmH<sub>2</sub>O.s/L)** | 214 | -1.61 ± 1.21 | 89 | -1.57 ± 1.07 | 393 | -1.55 ± 1.17 |
| **Food allergy (1 year)** | 318 | 7 (2.2%) | 121 | 32 (26.4%) | 689 | 53 (7.7%) |
| **Asthma (4 years)** | 333 | 40 (12.0%) | 133 | 42 (31.6%) | 761 | 117 (15.4%) |
| **Wheeze (4 years)** | 331 | 56 (16.9%) | 134 | 46 (34.3%) | 767 | 161 (21.0%) |
| **Eczema (4 years)** | 318 | 38 (11.9%) | 122 | 32 (26.2%) | 729 | 99 (13.6%) |
| **Rhinitis (4 years)** | 330 | 27 (8.2%) | 133 | 31 (23.3%) | 695 | 87 (12.5%) |

Abbreviations: SEIFA: Socio-Economic Indexes for Areas

**Table 2.** Descriptive characteristics of the Canadian Healthy Infant Longitudinal Development Study.

|  | Atopy subcohort (N = 937) |  |  |  | Study sample (N = 993) |  |
| --- | --- | --- | --- | --- | --- | --- |
|  | Non-atopic (N = 814) |  | Atopic (N = 123) |  |  |  |
|  | N | n (%)<br>mean ± SD | N | n (%)<br>mean ± SD | N | n (%)<br>mean ± SD |
| **Centre of recruitment** | 814 |  | 123 |  | 993 |  |
| Edmonton |  | 148 (18.2%) |  | 27 (22.0%) |  | 180 (18.1%) |
| Toronto |  | 122 (15.0%) |  | 24 (19.5%) |  | 172 (17.3%) |
| Vancouver |  | 227 (27.9%) |  | 51 (41.5%) |  | 296 (29.8%) |
| Winnipeg |  | 317 (38.9%) |  | 21 (17.1%) |  | 345 (34.7%) |
| **Year of birth** | 814 |  | 123 |  | 993 |  |
| 2009-2010 |  | 378 (46.4%) |  | 62 (50.4%) |  | 461 (46.4%) |
| 2011 |  | 436 (53.6%) |  | 61 (49.6%) |  | 532 (53.6%) |
| **Season of birth** | 814 |  | 123 |  | 993 |  |
| Autumn |  | 162 (19.9%) |  | 22 (17.9%) |  | 201 (20.2%) |
| Spring |  | 246 (30.2%) |  | 28 (22.8%) |  | 286 (28.8%) |
| Summer |  | 247 (30.3%) |  | 46 (37.4%) |  | 310 (31.2%) |
| Winter |  | 159 (19.5%) |  | 27 (22.0%) |  | 196 (19.7%) |
| **Male** | 814 | 450 (55.3%) | 123 | 73 (59.3%) | 993 | 550 (55.4%) |
| **Birthweight** | 789 | 3.46 ± 0.49 | 120 | 3.45 ± 0.44 | 964 | 3.5 ± 0.5 |
| **Breastfeeding duration (weeks)** | 814 | 44.36 ± 26.69 | 123 | 41.78 ± 26.73 | 993 | 44.3 ± 26.6 |
| **Maternal age at conception (years)** | 795 | 32.16 ± 4.41 | 121 | 33.25 ± 4.80 | 971 | 32.3 ± 4.5 |
| **Lone parenthood** | 797 | 40 (5.0%) | 122 | 5 (4.1%) | 975 | 46 (4.7%) |
| **Child ethnicity** | 814 |  | 123 |  | 993 |  |
| Caucasian/white |  | 608 (74.7%) |  | 64 (52.0%) |  | 704 (70.9%) |
| Multiracial |  | 116 (14.3%) |  | 26 (21.1%) |  | 153 (15.4%) |
| Other |  | 90 (11.1%) |  | 33 (26.8%) |  | 136 (13.7%) |
| **Caregiver university educated** | 814 | 669 (82.2%) | 123 | 97 (78.9%) | 993 | 818 (82.4%) |
| **Household income** | 725 |  | 106 |  | 879 |  |
| < 80,000 CAD |  | 238 (32.8%) |  | 33 (31.1%) |  | 280 (31.9%) |
| 80,000 – 149,999 CAD |  | 322 (44.4%) |  | 45 (42.5%) |  | 388 (44.1%) |
| ≥ 150,000 CAD |  | 165 (22.8%) |  | 28 (26.4%) |  | 211 (24.0%) |
| **Family history of asthma** | 809 | 167 (20.6%) | 123 | 38 (30.9%) | 988 | 218 (22.1%) |
| **Maternal smoke exposure during pregnancy** | 798 | 133 (16.7%) | 122 | 18 (14.8%) | 976 | 155 (15.9%) |
| **Smoking inside the house (5 years)** | 644 | 5 (0.8%) | 101 | 0 (0.0%) | 792 | 6 (0.8%) |
| **House renovations (5 years)** | 794 | 260 (32.7%) | 116 | 31 (26.7%) | 965 | 305 (31.6%) |
| **Household size (4+ vs < 4)** | 794 | 440 (55.4%) | 116 | 74 (63.8%) | 965 | 541 (56.1%) |
| **Pet ownership (5 years)** | 696 | 289 (41.5%) | 102 | 37 (36.3%) | 848 | 339 (40.0%) |
| **Functional residual capacity (litres)** | 43 | 0.67 ± 0.15 | 5 | 0.77 ± 0.14 | 54 | 0.67 ± 0.15 |
| **Lung clearance index (turnovers)** | 43 | 7.37 ± 0.97 | 5 | 6.77 ± 0.29 | 54 | 7.27 ± 0.94 |
| **Resistance at 11Hz (cmH<sub>2</sub>O.s/L)** | 111 | 10.44±2.13 | 23 | 10.59±2.77 | 154 | 10.50 ± 2.25 |
| **Reactance at 11Hz (cmH<sub>2</sub>O.s/L)** | 111 | -2.56±1.64 | 23 | -2.74±1.56 | 154 | -2.66 ± 1.64 |
| **Asthma (5 years)** | 814 | 44 (5.4%) | 123 | 15 (12.2%) | 976 | 62 (6.4%) |
| **Wheeze (5 years)** | 697 | 58 (8.3%) | 110 | 17 (15.5%) | 857 | 79 (9.2%) |
| **Eczema (5 years)** | 698 | 97 (13.9%) | 109 | 28 (25.7%) | 857 | 132 (15.4%) |
| **Rhinitis (5 years)** | 814 | 44 (5.4%) | 123 | 28 (22.8%) | 976 | 77 (7.9%) |

Relative to body weight, children in CHILD had higher estimated daily intakes than pregnant women in BIS, except for DEP (Table 3). In both cohorts, the exposure distributions did not differ substantially between atopic and non-atopic children.

**Table 3.**
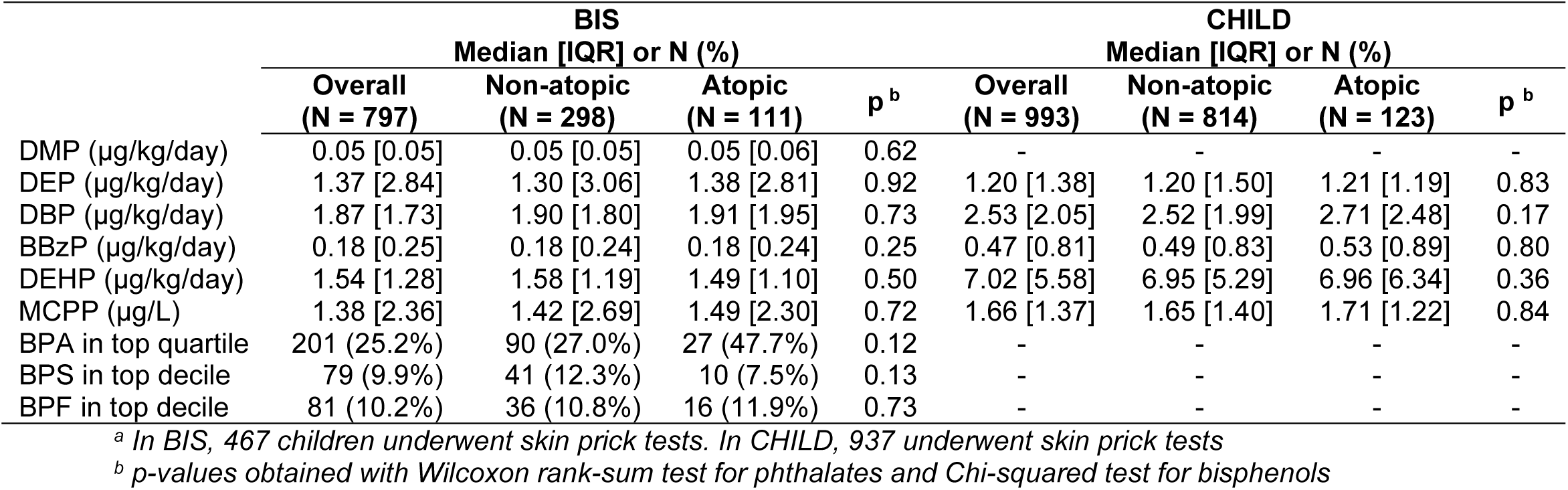
Exposure distributions stratified by atopic status ^a^.

### 3.2. Atopic and non-atopic phenotypes

In BIS, prenatal exposure to BBzP was inversely associated with atopy at age four (RR = 0.87; 95% CI: 0.79–0.97; Table S6). In contrast, no associations were observed between postnatal phthalate exposure and atopy at age five in the CHILD cohort.

Prenatal exposure to BPA in BIS was inversely associated with wheeze (RR = 0.64; 95% CI: 0.44– 0.94), but no other associations were found with overall outcomes at age four (Table 4). In CHILD, postnatal DBP was associated with an increased risk of asthma at age five (RR = 1.40; 95% CI: 1.10–1.77). Postnatal DEP (RR = 1.20; 95%CI: 1.07–1.35) and DBP (RR = 1.40; 95% CI: 1.11–1.76) were also associated with a higher risk of wheezing. The overall postnatal phthalate mixture was positively associated with asthma (RR = 1.69; 95% CI: 1.20–2.39) and wheeze (RR = 1.53; 95% CI: 1.11–2.10), with DEP and DBP identified as the most influential components (Figure S4). In addition, postnatal BBzP was associated with eczema (RR = 1.15; 95% CI: 1.03–1.28). Crude estimates are shown in Table S7.

**Table 4.**
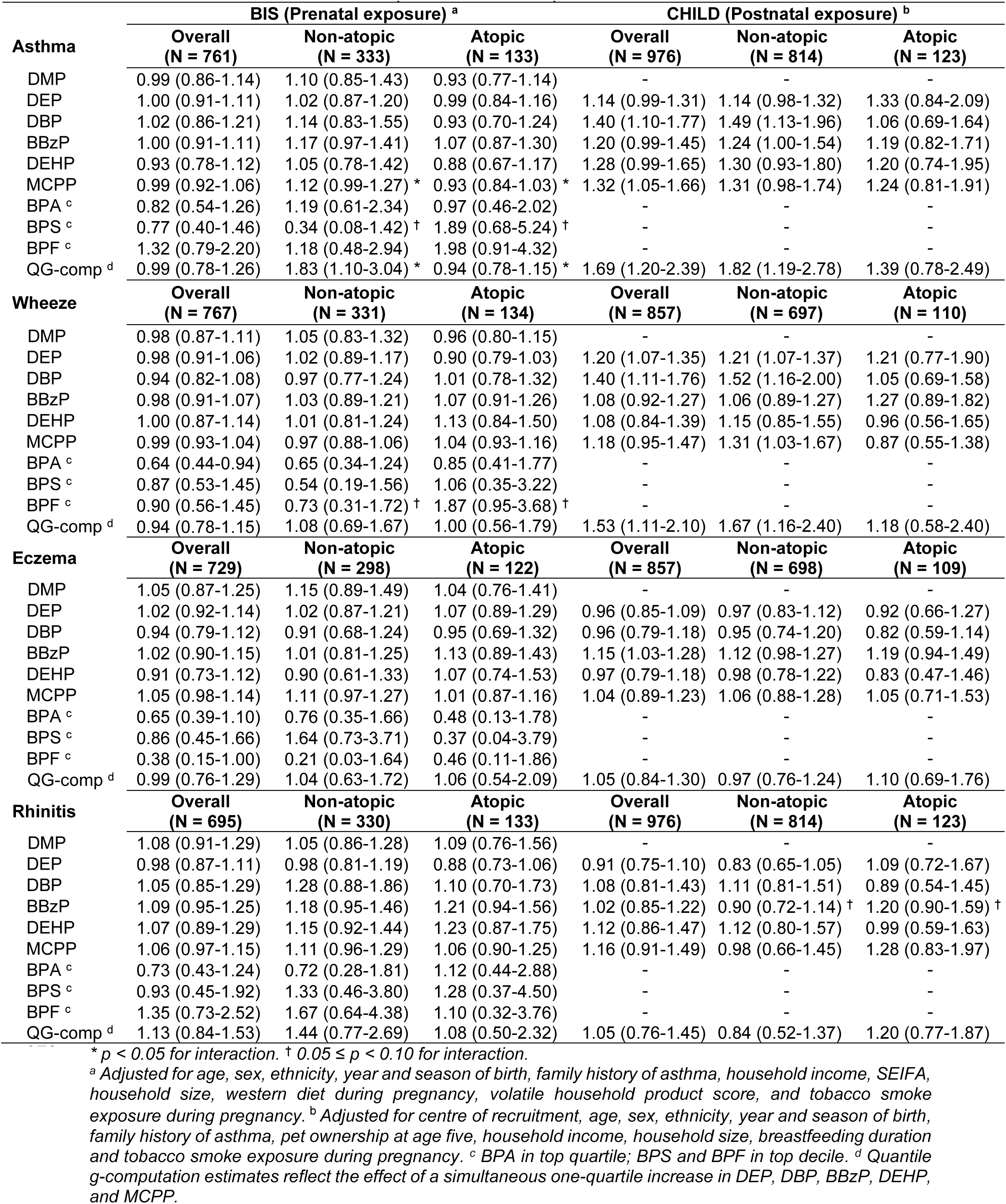
Adjusted risk ratios (and 95% confidence intervals) for outcomes associated with phthalates and bisphenols.

There was some evidence of effect measure modification by atopy in BIS. The association between prenatal MCPP levels and asthma differed by atopic status (p = 0.02 for interaction), with stronger estimates observed for non-atopic asthma (RR = 1.12; 95% CI: 0.99–1.27) than for atopic asthma (RR = 0.93; 95% CI: 0.84–1.03). Similarly, the prenatal phthalate mixture was more strongly associated with non-atopic asthma (RR = 1.83; 95% CI: 1.10–3.04) than with atopic asthma (RR = 0.94; 95% CI: 0.78–1.15; p = 0.02 for interaction). In contrast, prenatal BPS showed a stronger association with atopic asthma (RR = 1.89; 95% CI: 0.68–5.24; p = 0.05 for interaction), while prenatal BPF showed a stronger association with atopic wheeze (RR = 1.87; 95% CI: 0.95–3.68; p = 0.08 for interaction).

In the CHILD cohort, postnatal exposure to DBP (RR = 1.49; 95% CI: 1.13–1.96), BBzP (RR = 1.24; 95% CI: 1.00–1.54), and the overall phthalate mixture (RR = 1.82; 95% CI: 1.19–2.78) was associated with an increased risk of non-atopic asthma, but not atopic asthma. However, these associations did not differ substantially from those observed for atopic asthma (p for interaction = 0.19 for DBP, 0.83 for BBzP, 0.45 for the mixture). A similar pattern was observed for non-atopic wheeze, with positive associations for DEP (RR = 1.21; 95% CI, 1.07–1.37; p = 0.99 for interaction), DBP (RR = 1.52; 95% CI, 1.16–2.00; p = 0.14 for interaction), MCPP (RR = 1.31; 95% CI, 1.03–1.67; p = 0.12 for interaction) and the overall phthalate mixture (RR = 1.67; 95% CI: 1.16–2.40, p = 0.38 for interaction).

### 3.3. Supplementary analysis

Adjustment for additional covariates did not substantially change the estimates from the main analysis (Table S8). Results from BKMR were generally consistent with quantile G-computation (Figure S5-S6). However, prenatal and postnatal phthalate mixtures exhibited non-linear associations with atopic asthma. Specifically, the prenatal phthalate mixture demonstrated a U-shaped dose-response, while the postnatal mixture followed an inverted U-shaped relationship. The non-linear association observed with prenatal exposure was mainly driven by DEP, which was identified in our spline analyses (Figure S3).

In BIS, prenatal BPA was associated with a reduced risk of food allergy at one year of age (Table S9). Prenatal DMP exposure was inversely associated with sensitisation to both house dust mite and grass pollen, whereas postnatal MCCP showed a positive association (RR = 2.31; 95% CI: 1.11– 4.85; Table S10).

Regarding lung function outcomes, the prenatal phthalate mixture in BIS, was associated with increased FRC in atopic children (β = 0.27 SD; 95% CI: 0.02, 0.53), but tended to reduce FRC in non-atopic children (p = 0.003 for interaction; Table S11). The mixture was also associated with reduced resistance exclusively in atopic children (β =-0.34 SD; 95% CI:-0.58,-0.09; p = 0.02 for interaction). Additionally, prenatal BPF exposure tended to increase airway resistance in atopic children and decrease resistance in non-atopic children (p = 0.04 for interaction). Prenatal DEP exposure in BIS was associated with decreased resistance (p < 0.001 for interaction) and increased reactance only among atopic children (p = 0.04 for interaction), although spline models suggested non-linear dose–response relationships (Figure S3).

## 4. DISCUSSION

Prenatal and early childhood urinary concentrations of phthalate metabolites, measured in two separate cohorts, were associated with an increased risk of asthma in preschool-aged children. Associations between phthalate mixtures and asthma varied by atopic status across both exposure windows: mixtures showed a linear relationship with non-atopic asthma and non-linear associations with atopic asthma. Furthermore, prenatal exposure to BPF showed stronger associations with wheeze and airway resistance in atopic children, compared to non-atopic children. There was limited evidence of associations for other allergic outcomes. Overall, this study suggests that early-life exposure to phthalates may differentially influence asthma phenotypes, with little evidence of consistent associations with other allergic outcomes.

Few studies have investigated whether phthalate exposure may differentially influence atopic versus non-atopic phenotypes of childhood allergic conditions. Two European birth cohorts reported associations between prenatal DEHP metabolites and increased risk of atopic eczema, only in boys (10, 51). Differences in outcome definitions may explain the lack of replication in our study. Those studies assessed eczema over a broader age range (from birth to 4–5 years), whereas we assessed outcomes at a single time point. Furthermore, atopy was defined using total IgE, whereas we used SPT, which generally shows better diagnostic performance for early childhood allergies (52) Although we did not stratify our analyses by sex, future studies should investigate potential sex-specific susceptibilities for IgE-mediated outcomes. Our results for non-atopic asthma are in line with findings from the ALSPAC cohort, which reported that higher use of household chemical products during pregnancy, many of which contain phthalates (20, 53), was associated with an increased risk of non-atopic wheeze, but not atopic wheeze (54). Furthermore, other environmental chemicals, such as poly-and perfluoroalkyl substances and polycyclic aromatic hydrocarbons, have been associated with non-atopic asthma in children (55, 56). This supports the hypothesis that early-life chemical exposure may contribute to airway inflammation independent of IgE-mediated mechanisms.

There is also evidence suggesting that phthalates may induce a mixed inflammatory response. The Hokkaido study found that phthalate exposure in children aged 9–12 years was associated with both type 2 (eosinophilic) and non-type 2 (neutrophilic) inflammation biomarkers (57). Similarly, a Taiwanese adult case-control study reported associations between DEHP metabolites and both atopic and non-atopic asthma (58). In our study, associations between postnatal exposure to individual phthalates and asthma did not differ substantially by atopic status. Atopic asthma is often driven by Th2-mediated eosinophilic inflammation, whereas non-atopic asthma is more often associated with Th17-dominant neutrophilic inflammation (59). Animal studies have shown that phthalates can activate the p38 MAPK signalling pathway, leading to increased oxidative stress and a mixed Th2/Th17 immune response (60). This pathway is negatively regulated by estrogen receptor beta (ER_β_), which exerts anti-proliferative effects on airway smooth muscle (61). Given that phthalates are estrogenic and anti-estrogenic xenobiotics (62, 63), these compounds may disrupt ER_β_ signalling, leading to p38 MAPK activation and subsequent mixed Th2/Th17-mediated airway inflammation.

The distinct dose-response patterns observed for atopic versus non-atopic asthma, however, suggest potentially different mechanisms between phenotypes. The U-shaped association for atopic asthma is consistent with the non-linear dose-response relationships that are possible for endocrine-disrupting chemicals (64). The Maternal-Infant Research on Environmental Chemicals Study, for instance, reported an inverted U-shaped association between maternal MCPP concentrations and cord blood IgE levels (65). These findings support the notion that phthalates may influence immune development through nonlinear effects. Non-monotonic dose-response patterns have been described in hormonal pathways affected by phthalates (64), such as estrogen and androgen signalling, which may explain the male-specific associations with atopic asthma observed in earlier studies (10, 51). In contrast, the linear associations we observed for non-atopic asthma may reflect a dose-dependent increase in oxidative stress (66), which has been implicated in Th17 neutrophilic inflammation (67).

Due to the low detection frequencies of maternal bisphenols, our ability to assess their associations with respiratory and allergic outcomes was limited. Nonetheless, we found that prenatal exposure to BPF was more strongly associated with atopic wheeze and increased airway resistance in atopic children. Although mechanistic evidence specific to BPF and atopic outcomes remains limited, BPF has estrogenic activity (68), which may promote a Th2 immune response (69). Experimental studies have also shown that gestational and lactational exposure to BPF can induce airway eosinophilic inflammation and cellular infiltration in female offspring, potentially through disruption of redox homeostasis and immunosuppressive pathways (70).

We also found that prenatal exposure to phthalate mixtures was associated with a lower FRC in non-atopic children, consistent with a paradigm of lower lung volumes predisposing to symptomatic lung disease in non-atopic children (71). In contrast, increased FRC in atopic children may indicate air trapping due to small airway obstruction (71). However, these findings need to be confirmed with more sensitive methods for assessing static lung volumes, such as whole-body plethysmography. Although the observed reductions in airway resistance associated with prenatal phthalate mixture exposure among atopic children seem contradictory, the presence of non-monotonic dose-response relationships observed in this study suggests heterogeneous effects across exposure levels. Further studies are needed to understand how early-life chemical exposure influences respiratory mechanics in developing lungs.

The inverse associations observed between prenatal exposure to phthalates or bisphenols and allergic outcomes, including food allergy, are unlikely to be causal. There is no biological basis to suggest that prenatal phthalates and bisphenols protect against allergic conditions. These findings could be the result of unmeasured confounding or chance findings. Replication in larger cohorts is warranted to clarify these associations.

This study has important strengths. We examined both prenatal and postnatal exposure windows, and used objective outcome measures, including skin prick testing, lung function assessments, and oral food challenge, which is the gold standard for diagnosing IgE-mediated food allergies (52). The inclusion of two population-based birth cohorts enhances the generalisability of our findings to high-income, English-speaking settings. However, several limitations should be acknowledged. First, phthalates and bisphenols have short biological half-lives and are eliminated within 24–48 hours (72). Thus, single-spot urine samples may not accurately reflect long-term exposure and could result in nondifferential misclassification, biasing estimates toward the null. In contrast, childhood measurements were taken at widely spaced intervals, and we averaged concentrations across three time points to better represent overall early-childhood exposure. However, this approach may have hidden specific windows of vulnerability. Furthermore, the lack of time-of-day data for urine collection in CHILD may have introduced further misclassification. Second, differences in outcome ascertainment between cohorts limited direct comparability. In BIS, allergic outcomes were primarily parent-reported and susceptible to overreporting and non-differential outcome misclassification bias. In contrast, CHILD relied on physician diagnoses, which may explain the lower prevalence of allergic outcomes. A smaller proportion of atopic children in CHILD may have limited statistical power to detect modest associations, particularly in subgroups related to lung function. Third, while skin prick testing provides an objective measure of sensitisation, it may not capture the full heterogeneity of allergic phenotypes. For example, some children with a local airway Th2-immune response may test negative on SPT despite underlying eosinophilic inflammation (73). Future studies should consider more granular phenotyping approaches, such as molecular analysis of nasal epitheliums or induced sputum. Finally, lung function measurements were conducted when children were asymptomatic, which may have underestimated the presence or severity of airway dysfunction associated with environmental exposures.

In summary, exposure to phthalates during late pregnancy and early childhood may differentially influence asthma phenotypes at preschool age, with limited evidence for consistent associations with other allergic outcomes. Future studies should incorporate more granular phenotyping approaches to characterise underlying mechanisms. In addition, greater harmonisation of outcome definitions across cohorts is needed to enable direct comparisons and strengthen causal inference in future studies.

## CRediT authorship contribution statement

**Thomas Boissiere-O’Neill**: Conceptualisation, Methodology, Formal analysis Writing – original draft. **Nina Lazarevic**: Conceptualisation, Methodology, Writing – review and editing. **Anne-Louise Ponsonby**: Conceptualisation, Methodology, Writing – review and editing. **Peter D. Sly**: Conceptualisation, Writing – review and editing. **Aimin Chen**: Conceptualisation, Writing – review and editing. **Tamara L. Blake**: Writing – review and editing. **Jeffrey R. Brook**: Writing – review and editing. **Cassidy Du Berry**: Validation, Writing – review and editing. **Louise King**: Validation, Writing – review and editing**. Piushkumar J. Mandhane**: Writing – review and editing. **Theo J. Moraes**: Writing – review and editing. **Elinor Simons**: Writing – review and editing. **Padmaja Subbarao**: Writing – review and editing. **Dwan Vilcins**: Supervision, Conceptualisation, Methodology, Writing – review and editing.

## Data statement

The data that support the findings of this study are not openly available due to reasons of sensitivity and are available from childstudy.ca and barwoninfantstudy.org.au

## Acknowledgements

We thank the CHILD Cohort Study (CHILD) participant families for their dedication and commitment to advancing health research. CHILD was initially funded by CIHR and AllerGen NCE. Visit CHILD at childstudy.ca. We thank the BIS participants for the generous contribution they have made to this project. We also thank current and past staff for their efforts in recruiting and maintaining the cohort and in obtaining and processing the data and biospecimens. The establishment work and infrastructure for the BIS was provided by the Murdoch Children’s Research Institute, Deakin University and Barwon Health. Subsequent funding was secured from the National Health and Medical Research Council of Australia, The Jack Brockhoff Foundation, the Scobie Trust, the Shane O’Brien Memorial Asthma Foundation, the OurWomen’s Our Children’s Fund Raising Committee Barwon Health, The Shepherd Foundation, the Rotary Club of Geelong, the Ilhan Food Allergy Foundation, GMHBA Ltd, The Gandel Foundation, The Percy Baxter Charitable Trust, Perpetual Trustees and the Gwenyth Raymond Trust. Vanguard Investments Australia Ltd provided funding for analysis of plasticizers in biospecimens. In-kind support was provided by the Cotton On Foundation and CreativeForce. Research at Murdoch Children’s Research Institute is supported by the Victorian Government’s Operational Infrastructure Support Program.

