## Supplementary materials for "Phthalates, Bisphenols, and Childhood Allergic Phenotypes: Findings from Two Birth Cohort Studies"

### Table S1. Distribution of urinary phthalate and bisphenol metabolites across cohorts (µg/L)

|  | Detection frequency | GM (GSD) | Q10 | Q25 | Q50 | Q75 | Q90 | Maximum |
| --- | --- | --- | --- | --- | --- | --- | --- | --- |
| MMP (BIS) | 98.6% | 1.57 (2.40) | 0.68 | 0.99 | 1.53 | 2.47 | 4.14 | 41.66 |
| MEP |  |  |  |  |  |  |  |  |
| BIS | 99.4% | 47.12 (3.72) | 11.10 | 20.52 | 39.55 | 95.80 | 271.64 | 5040.50 |
| CHILD |  |  |  |  |  |  |  |  |
| 3 months | 92.3% | 7.62 (3.11) | 2.18 | 4.01 | 7.08 | 13.56 | 28.59 | 4584 |
| 12 months | 97.2% | 12.37 (3.30) | 3.60 | 6.09 | 11.40 | 22.09 | 45.49 | 5210 |
| 36 months | 95.6% | 13.94 (3.01) | 3.96 | 7.12 | 12.60 | 26.25 | 51.96 | 1804.09 |
| ΣMBP |  |  |  |  |  |  |  |  |
| BIS | 91.3% | 51.49 (1.93) | 23.39 | 33.87 | 51.73 | 76.26 | 117.42 | 713.67 |
| CHILD |  |  |  |  |  |  |  |  |
| 3 months | 99.5% | 15.06 (2.29) | 5.77 | 8.89 | 14.49 | 24.40 | 41.86 | 563.2 |
| 12 months | 99.7% | 24.36 (2.20) | 10.62 | 14.87 | 22.88 | 37.61 | 66.71 | 1002.5 |
| 36 months | 99.8% | 32.04 (2.24) | 13.91 | 21.04 | 31.68 | 50.75 | 76.76 | 1144.14 |
| MBzP |  |  |  |  |  |  |  |  |
| BIS | 98.0% | 5.30 (2.94) | 1.51 | 2.75 | 5.33 | 9.94 | 19.85 | 171.70 |
| CHILD |  |  |  |  |  |  |  |  |
| 3 months | 69.3% | 1.81 (3.29) | 0.41 | 0.82 | 1.85 | 3.86 | 7.87 | 222 |
| 12 months | 87.8% | 5.57 (3.76) | 1.13 | 2.26 | 4.96 | 12.78 | 32.16 | 328.889 |
| 36 months | 93.4% | 8.48 (3.44) | 2.07 | 3.90 | 7.56 | 18.17 | 42.92 | 1071 |
| MEHP (CHILD) |  |  |  |  |  |  |  |  |
| 3 months | 37.4% | 0.52 (3.98) | – | – | – | 1.26 | 2.70 | 59.3333 |
| 12 months | 84.0% | 1.77 (2.41) | 0.66 | 1.06 | 1.73 | 2.88 | 5.04 | 96.3 |
| 36 months | 87.7% | 2.23 (2.36) | 0.92 | 1.42 | 2.21 | 3.57 | 5.64 | 142.8 |
| MEHHP |  |  |  |  |  |  |  |  |
| BIS | 99.9% | 11.26 (2.20) | 4.76 | 7.48 | 11.29 | 16.54 | 25.35 | 3618.85 |
| CHILD |  |  |  |  |  |  |  |  |
| 3 months | 69.4% | 1.55 (2.84) | – | – | 1.64 | 2.99 | 5.36 | 47.8 |
| 12 months | 97.6% | 7.78 (2.37) | 2.95 | 4.60 | 7.78 | 13.34 | 21.20 | 361 |
| 36 months | 99.2% | 11.48 (2.37) | 4.62 | 7.17 | 11.17 | 18.08 | 30.25 | 284.667 |
| MEOHP |  |  |  |  |  |  |  |  |
| BIS | 100.0% | 9.13 (2.13) | 3.84 | 6.17 | 8.94 | 13.44 | 20.09 | 1729.21 |
| CHILD |  |  |  |  |  |  |  |  |
| 3 months | 65.3% | 1.30 (2.67) | – | – | 1.41 | 2.47 | 4.21 | 31.28 |
| 12 months | 96.0% | 5.48 (2.32) | 2.05 | 3.29 | 5.40 | 9.69 | 14.77 | 248 |
| 36 months | 99.0% | 8.21 (2.31) | 3.47 | 5.11 | 8.15 | 12.43 | 20.84 | 233.1 |
| MECPP (BIS) | 100.0% | 13.70 (1.99) | 6.37 | 9.22 | 13.07 | 19.28 | 29.07 | 2107.17 |
| MCPP |  |  |  |  |  |  |  |  |
| BIS | 79.9% | 1.00 (5.99) | – | 0.53 | 1.39 | 2.94 | 6.58 | 284.24 |
| CHILD |  |  |  |  |  |  |  |  |
| 3 months | 43.4% | 0.58 (2.60) | – | – | – | 1.12 | 1.83 | 35.0667 |
| 12 months | 84.9% | 1.91 (2.22) | – | 1.13 | 1.87 | 3.03 | 5.10 | 115.5 |
| 36 months | 81.3% | 1.89 (2.84) | – | 1.08 | 2.03 | 3.41 | 5.93 | 170.52 |
| BPA (BIS) | 53.7% | 0.65 (4.00) | – | – | – | 2.17 | 4.15 | 57.06 |
| BPS (BIS) | 34.8% | 0.05 (2.80) | – | – | – | – | 0.23 | 54.98 |
| BPF (BIS) | 15.7% | 0.24 (2.15) | – | – | – | – | 0.72 | 22.67 |

*Abbreviations: BIS: Barwon Infant Study; BPA: Bisphenol A; BPF: Bisphenol F; BPS: Bisphenol S; CHILD: Canadian Healthy Infant Longitudinal Development Study; GM: Geometric mean; GSD: Geometric standard deviation; MBP: mono-benzyl phthalate; MBzP: Mono-benzyl phthalate; MCPP: Mono(3-carboxypropyl) phthalate;; MECPP: Mono(2-ethyl-5-carboxypentyl) phthalate; MEHHP: Mono(2-ethyl-5-hydroxyhexyl) phthalate; MEHP: Mono-2-ethylhexyl phthalate; MEOHP: Mono(2-ethyl-5-oxohexyl) phthalate; MEP: mono-ethyl phthalate; MMP: Mono-methyl phthalate.*

### Table S2. List of allergens used in skin prick testing in both cohorts

| **Category** | **Allergen** | **BIS**  **(4 years)** | **CHILD**  **(5 years)** |
| --- | --- | --- | --- |
| Outdoor  aeroallergens | Grass | ✔ (ryegrass) | ✔ (grass mix) |
|  | Midwest trees |  | ✔ |
|  | Weed mix |  | ✔ |
|  | Mixed ragweed |  | ✔ |
| House dust mites | *D. pteronyssinus* | ✔ | ✔ |
|  | *D. farinae* |  | ✔ |
| Pets | Dog | ✔ | ✔ |
|  | Cat | ✔ | ✔ |
| Fungi | Alternaria tenuis | ✔ | ✔ |
|  | Cladosporium |  | ✔ |
|  | Penicillium mixed |  | ✔ |
|  | Aspergillus fumigatus |  | ✔ |
| Pests | German cockroach |  | ✔ |
| Food | Sesame | ✔ |  |
|  | Cashew | ✔ |  |
|  | Peanut | ✔ | ✔ |
|  | Egg | ✔ | ✔ |
|  | Cow milk | ✔ | ✔ |
|  | Soybean |  | ✔ |

### Table S3. Covariate definitions and availability

| Covariate | Definition | Variable availability | | |
| --- | --- | --- | --- | --- |
|  |  | BIS | | CHILD |
| **Sociodemographic factors** | | |  | |
| Maternal age at conception | Continuous (years) | ✔ | | ✔ |
| Child ethnicity | Categorical (Caucasian/white; other) | ✔ | | ✔ |
| Marital status | Categorical (single/not married; other) | ✔ | | ✔ |
| Remoteness area | Categorical (major city; inner regional) | ✔ | |  |
| Parental education | Categorical (University educated; other) | ✔ | | ✔ |
| Household income | Categorical | ✔ | | ✔ |
| Socio-Economic Indexes for Areas (SEIFA) | An Australian composite measure of area-based socioeconomic status. SEIFA scores were stratified into tertiles, with lower values denoting areas of greater disadvantage and higher values indicating areas of greater advantage (1) | ✔ | |  |
| **Perinatal and Birth Characteristics** | | |  | |
| Year of birth | Categorical | ✔ | | ✔ |
| Season of birth | Categorical (spring, summer, autumn, winter) | ✔ | | ✔ |
| Birthweight | Continuous (kilograms) | ✔ | | ✔ |
| Sex | Binary (male; female) | ✔ | | ✔ |
| Any breastfeeding duration | Continuous (weeks) | ✔ | | ✔ |
| **Family and Environmental Factors** | | |  | |
| Family history of asthma | History of asthma in a 1^st^-degree relative (yes; no) | ✔ | | ✔ |
| Maternal smoke exposure | Any tobacco smoke exposure during pregnancy (yes; no) | ✔ | | ✔ |
| Smoking inside the house | Any active smoking inside the house (binary) during the four-year (BIS) or five-year review (CHILD) | ✔ | | ✔ |
| Household renovations | Household renovation (binary) during pregnancy (BIS) or at the 5-year review (CHILD) | ✔ | | ✔ |
| Household size | Categorical (BIS: 2, 3, ≥ 4; CHILD: <4, 4+) | ✔ | | ✔ |
| Pet ownership | Any pet ownership (binary) during pregnancy (BIS) or at the 5-year review (CHILD) | ✔ | | ✔ |
| Maternal diet during pregnancy | Previously assessed using a food frequency questionnaire and principal component analysis (2, 3). Three dietary patterns were identified: (i) modern healthy, (ii) western unhealthy, and (iii) traditional Anglo-Saxon. Factor loadings were categorised into tertiles, reflecting diet adherence | ✔ | |  |
| Chemical burden score | Composite measure of the sum of volatile household product use frequencies during pregnancy (4) | ✔ | |  |
| **Other** |  |  | |  |
| Centre of recruitment | Categorical (Vancouver, Toronto, Edmonton, Winnipeg) |  | | ✔ |
| Age at outcome assessment | Continuous (years) | ✔ | | ✔ |

Abbreviations: *BIS: Barwon Infant Study; CHILD: Canadian Healthy Infant Longitudinal Development Study*

### Table S4. Exposure distributions in study sample vs full exposure data

|  | BIS | | CHILD | |
| --- | --- | --- | --- | --- |
|  | Sample with exposure data  (N = 847) | Study sample  (N = 797) | Sample with exposure data  (N = 1,075) | Study sample  (N = 993) |
| DMP (µg/kg/day) | 0.05 [0.05] | 0.05 [0.05] | – | – |
| DEP (µg/kg/day) | 1.37 [2.69] | 1.29 [2.84] | 1.20 [1.36] | 1.20 [1.38] |
| DBP (µg/kg/day) | 1.86 [1.73] | 1.87 [1.73] | 2.53 [2.06] | 2.53 [2.05] |
| BBzP (µg/kg/day) | 0.18 [0.25] | 0.18 [0.25] | 0.47 [0.81] | 0.47 [0.81] |
| DEHP (µg/kg/day) | 1.54 [1.27] | 1.54 [1.28] | 6.95 [5.42] | 7.02 [5.58] |
| MCPP (µg/L) | 1.39 [2.42] | 1.38 [2.36] | 1.65 [1.37] | 1.66 [1.37] |
| BPA in top quartile | 212 (25.0%) | 201 (25.2%) | – | – |
| BPS in top decile | 85 (10.0%) | 79 (9.9%) | – | – |
| BPF in top decile | 85 (10.0%) | 81 (10.2%) | – | – |

*Values shown as median [IQR] or n(%).*

*Abbreviations: BBzP = butylbenzyl phthalate; BIS: Barwon Infant study; BPA: bisphenol A; BPF: bisphenol F; BPS: bisphenol S; CHILD: Canadian Healthy Infant Longitudinal Development study; DBP: dibutyl phthalate; DEP: diethyl phthalate; DEHP = di(2-ethylhexyl) phthalate; DMP: dimethyl phthalate; MCPP = mono(3-carboxypropyl) phthalate.*

### Table S5. Outcome prevalences in the study sample vs complete outcome data

|  | BIS | | CHILD | |
| --- | --- | --- | --- | --- |
|  | Sample with outcome data  (N = 951) | Study sample  (N = 797) | Sample with outcome data  (N = 2,720) | Study sample  (N = 993) |
| Asthma | 144 (16.1%) | 117 (15.4%) | 165 (6.2%) | 62 (6.4%) |
| Missing | 58 | 36 | 74 | 17 |
| Wheeze | 195 (21.5%) | 161 (21.0%) | 226 (9.8%) | 79 (9.2%) |
| Missing | 45 | 15 | 418 | 136 |
| Eczema | 103 (12.3%) | 99 (13.6%) | 357 (15.5%) | 132 (15.4%) |
| Missing | 114 | 46 | 415 | 136 |
| Rhinitis | 99 (12.4%) | 87 (12.5%) | 188 (7.1%) | 77 (7.9%) |
| Missing | 154 | 52 | 76 | 17 |
| Atopy | 144 (28.1%) | 134 (28.7%) | 305 (12.0%) | 123 (13.1%) |
| Missing | 438 | 108 | 186 | 56 |
| Food allergy | 58 (7.2%) | 53 (7.7%) | – | – |
| Missing | 147 | 108 | – | – |
| FRC (litres) | 0.57 ± 0.10 | 0.57 ± 0.10 | 0.68± 0.13 | 0.67 ± 0.15 |
| Missing | 729 | 262 | 2,515 | 939 |
| LCI_2.5_ (turnovers) | 6.24 ± 0.42 | 6.22 ± 0.42 | 7.33 ± 0.98 | 7.27 ± 0.94 |
| Missing | 729 | 262 | 2,515 | 939 |
| R10-11 (cmH_2_O.s/L) | 8.34 ± 1.92 | 8.27± 1.90 | 10.38 ±2.34 | 10.50 ± 2.25 |
| Missing | 519 | 404 | 2,220 | 839 |
| X10-11 (cmH_2_O.s/L) | -1.58 ± 1.17 | -1.55 ± 1.17 | -2.78 ± 1.55 | -2.66 ± 1.64 |
| Missing | 519 | 404 | 2,220 | 839 |

*Values shown as mean* ± SD or *n (%).*

*Abbreviations: BIS: Barwon Infant study; CHILD: Canadian Healthy Infant Longitudinal Development study; FRC: Functional residual capacity; LCI_2.5_: Lung clearance index; R: Resistance; X: Reactance.*

### Table S6. Risk ratios (and 95% confidence intervals) for atopy associated with phthalates and bisphenols

|  | **BIS (Prenatal exposure; N = 467) ^a^** | | | **CHILD (Postnatal exposure; N = 937) ^b^** | | |
| --- | --- | --- | --- | --- | --- | --- |
|  | **Crude** | **Minimally adjusted** | **Fully**  **adjusted** | **Crude** | **Minimally adjusted** | **Fully**  **adjusted** |
| DMP | 0.95 (0.86-1.06) | 0.94 (0.83-1.05) | 0.93 (0.83-1.06) | – | – | – |
| DEP | 0.98 (0.91-1.06) | 0.99 (0.91-1.08) | 1.00 (0.92-1.08) | 0.98 (0.87-1.09) | 0.98 (0.87-1.10) | 0.98 (0.87-1.10) |
| DBP | 0.99 (0.85-1.16) | 0.97 (0.83-1.14) | 0.97 (0.82-1.14) | 1.15 (0.94-1.40) | 1.14 (0.92-1.41) | 1.14 (0.92-1.42) |
| BBzP | 0.91 (0.83-1.01) | 0.87 (0.79-0.97) | 0.87 (0.78-0.96) | 1.01 (0.91-1.12) | 1.11 (0.98-1.25) | 1.11 (0.98-1.25) |
| DEHP | 0.90 (0.78-1.02) | 0.88 (0.76-1.02) | 0.88 (0.76-1.03) | 1.09 (0.90-1.31) | 1.07 (0.88-1.30) | 1.07 (0.88-1.30) |
| MCPP | 0.99 (0.94-1.04) | 0.98 (0.92-1.04) | 0.97 (0.92-1.03) | 1.00 (0.84-1.20) | 1.05 (0.87-1.27) | 1.04 (0.86-1.27) |
| BPA ^c^ | 0.75 (0.52-1.09) | 0.73 (0.51-1.06) | 0.74 (0.52-1.08) | – | – | – |
| BPS ^c^ | 0.66 (0.37-1.18) | 0.57 (0.32-1.04) | 0.57 (0.31-1.03) | – | – | – |
| BPF ^c^ | 1.08 (0.69-1.69) | 1.03 (0.66-1.63) | 1.11 (0.70-1.76) | – | – | – |
| QG-comp ^d^ | 0.96 (0.78-1.18) | 0.91 (0.75-1.12) | 0.92 (0.75-1.13) | 1.10 (0.88-1.38) | 1.19 (0.94-1.50) | 1.19 (0.95-1.50) |

*^a^ Minimally adjusted models were adjusted for age, sex, ethnicity, year and season of birth, family history of asthma, household income, SEIFA, household size, western diet during pregnancy, volatile household product score, and tobacco smoke exposure during pregnancy.* *Fully adjusted models were further adjusted for pet ownership during pregnancy, living in a major Australian city, and house renovations during pregnancy*

^b^ *Minimally adjusted models were adjusted for centre of recruitment, age, sex, ethnicity, year and season of birth, family history of asthma, household income, pet ownership at age five, and tobacco smoke exposure during pregnancy. Fully adjusted models were further adjusted for birthweight and house renovations at age five.*

*^c^ BPA in top quartile; BPS and BPF in top decile.*

*^d^ Quantile g-computation estimates reflect the effect of a simultaneous one-quartile increase in DEP, DBP, BBzP, DEHP, and MCPP.*

*Abbreviations: BBzP = butylbenzyl phthalate; BIS: Barwon Infant study; BPA: bisphenol A; BPF: bisphenol F; BPS: bisphenol S; CHILD: Canadian Healthy Infant Longitudinal Development study; DBP: dibutyl phthalate; DEP: diethyl phthalate; DEHP = di(2-ethylhexyl) phthalate; DMP: dimethyl phthalate; MCPP = mono(3-carboxypropyl) phthalate; QG-comp: quantile G-computation.*

### Table S7. Crude risk ratios (and 95% confidence intervals) for outcomes associated with phthalates and bisphenols

|  | **BIS (Prenatal exposure)** | | | **CHILD (Postnatal exposure)** | | |
| --- | --- | --- | --- | --- | --- | --- |
| **Asthma** | **Overall**  **(N = 761)** | **Non-atopic**  **(N = 333)** | **Atopic**  **(N = 133)** | **Overall**  **(N = 976)** | **Non-atopic**  **(N = 814)** | **Atopic**  **(N = 123)** |
| DMP | 1.00 (0.88-1.15) | 1.12 (0.85-1.47) | 0.93 (0.79-1.09) | – | – | – |
| DEP | 1.00 (0.91-1.10) | 1.02 (0.88-1.17) | 0.95 (0.82-1.10) | 1.14 (1.00-1.29) | 1.16 (1.01-1.33) | 1.18 (0.78-1.77) |
| DBP | 1.05 (0.88-1.25) | 1.19 (0.84-1.70) | 1.05 (0.82-1.34) | 1.39 (1.11-1.73) | 1.49 (1.13-1.95) | 1.04 (0.72-1.50) |
| BBzP | 1.01 (0.91-1.13) | 1.18 (0.98-1.41) | 1.08 (0.91-1.27) | 1.11 (0.96-1.29) | 1.16 (0.98-1.38) | 1.11 (0.83-1.48) |
| DEHP | 0.95 (0.80-1.13) | 1.05 (0.79-1.39) | 0.93 (0.72-1.21) | 1.20 (0.94-1.52) | 1.22 (0.90-1.66) | 1.12 (0.74-1.69) |
| MCPP | 1.00 (0.93-1.07) | 1.14 (1.01-1.28) * | 0.94 (0.85-1.03) * | 1.27 (1.03-1.57) | 1.29 (0.98-1.70) | 1.20 (0.81-1.78) |
| BPA ^a^ | 0.81 (0.54-1.22) | 1.16 (0.61-2.19) | 0.92 (0.47-1.80) | – | – | – |
| BPS ^a^ | 0.76 (0.40-1.45) | 0.37 (0.09-1.55) | 1.29 (0.53-3.14) | – | – | – |
| BPF ^a^ | 1.31 (0.80-2.14) | 1.18 (0.48-2.89) | 1.46 (0.76-2.82) | – | – | – |
| QG-comp ^b^ | 1.04 (0.81-1.33) | 1.85 (1.12-3.05) * | 0.81 (0.46-1.40) * | 1.55 (1.11-2.17) | 1.73 (1.12-2.67) | 1.18 (0.69-2.01) |
| **Wheeze** | **Overall**  **(N = 767)** | **Non-atopic**  **(N = 331)** | **Atopic**  **(N = 134)** | **Overall**  **(N = 857)** | **Non-atopic**  **(N = 697)** | **Atopic**  **(N = 110)** |
| DMP | 1.01 (0.90-1.13) | 1.07 (0.86-1.34) | 0.97 (0.83-1.14) | – | – | – |
| DEP | 0.99 (0.91-1.07) | 1.03 (0.90-1.17) | 0.90 (0.80-1.01) | 1.20 (1.08-1.34) | 1.23 (1.09-1.38) | 1.16 (0.82-1.66) |
| DBP | 0.96 (0.83-1.10) | 0.97 (0.76-1.24) | 1.12 (0.88-1.42) | 1.41 (1.16-1.72) | 1.51 (1.20-1.91) | 1.06 (0.74-1.52) |
| BBzP | 1.00 (0.92-1.09) | 1.08 (0.93-1.26) | 1.11 (0.95-1.28) | 1.07 (0.94-1.21) | 1.06 (0.92-1.23) | 1.20 (0.91-1.59) |
| DEHP | 1.02 (0.89-1.15) | 1.00 (0.81-1.24) | 1.17 (0.91-1.49) | 1.12 (0.89-1.41) | 1.18 (0.90-1.53) | 0.91 (0.56-1.48) |
| MCPP | 1.00 (0.95-1.05) | 0.99 (0.91-1.08) | 1.06 (0.95-1.17) | 1.19 (0.97-1.47) | 1.34 (1.05-1.71) | 0.91 (0.63-1.31) |
| BPA ^a^ | 0.65 (0.45-0.94) | 0.65 (0.35-1.22) | 0.83 (0.43-1.61) | – | – | – |
| BPS ^a^ | 0.87 (0.52-1.43) | 0.54 (0.20-1.46) | 0.87 (0.29-2.55) | – | – | – |
| BPF ^a^ | 0.96 (0.60-1.53) | 0.80 (0.33-1.93) | 1.55 (0.86-2.79) | – | – | – |
| QG-comp ^b^ | 1.00 (0.81-1.22) | 1.15 (0.75-1.77) | 1.08 (0.63-1.86) | 1.54 (1.14-2.07) | 1.74 (1.22-2.48) | 1.14 (0.60-2.17) |
| **Eczema** | **Overall**  **(N = 729)** | **Non-atopic**  **(N = 298)** | **Atopic**  **(N = 122)** | **Overall**  **(N = 857)** | **Non-atopic**  **(N = 698)** | **Atopic**  **(N = 109)** |
| DMP | 1.06 (0.91-1.23) | 1.17 (0.93-1.47) | 1.01 (0.80-1.26) | – | – | – |
| DEP | 1.02 (0.93-1.13) | 1.01 (0.86-1.19) | 1.05 (0.90-1.24) | 0.96 (0.85-1.09) | 0.96 (0.83-1.11) | 0.97 (0.69-1.37) |
| DBP | 0.95 (0.80-1.13) | 0.90 (0.67-1.22) | 0.93 (0.71-1.21) | 0.98 (0.82-1.17) | 0.96 (0.77-1.20) | 0.82 (0.61-1.11) |
| BBzP | 1.02 (0.91-1.15) | 1.00 (0.82-1.22) | 1.10 (0.89-1.35) | 1.15 (1.05-1.27) | 1.14 (1.02-1.28) | 1.17 (0.94-1.45) |
| DEHP | 0.93 (0.77-1.13) | 0.88 (0.60-1.27) | 1.08 (0.79-1.48) | 0.99 (0.82-1.18) | 0.99 (0.80-1.23) | 0.89 (0.57-1.39) |
| MCPP | 1.06 (0.99-1.14) | 1.11 (0.98-1.25) | 1.03 (0.91-1.17) | 1.04 (0.88-1.22) | 1.07 (0.88-1.30) | 1.00 (0.71-1.41) |
| BPA ^a^ | 0.61 (0.37-1.01) | 0.73 (0.35-1.55) | 0.45 (0.14-1.40) | – | – | – |
| BPS ^a^ | 0.93 (0.48-1.77) | 1.66 (0.77-3.58) | 0.46 (0.06-3.84) | – | – | – |
| BPF ^a^ | 0.37 (0.14-1.00) | 0.23 (0.03-1.76) | 0.48 (0.11-1.97) | – | – | – |
| QG-comp ^b^ | 1.03 (0.79-1.36) | 1.00 (0.60-1.67) | 1.12 (0.61-2.07) | 1.06 (0.85-1.30) | 1.00 (0.78-1.27) | 1.13 (0.73-1.76) |
| **Rhinitis** | **Overall**  **(N = 695)** | **Non-atopic**  **(N = 330)** | **Atopic**  **(N = 133)** | **Overall**  **(N = 976)** | **Non-atopic**  **(N = 814)** | **Atopic**  **(N = 123)** |
| DMP | 1.12 (0.94-1.33) | 1.07 (0.88-1.30) | 1.11 (0.78-1.58) | – | – | – |
| DEP | 0.99 (0.89-1.11) | 1.01 (0.84-1.21) | 0.92 (0.78-1.07) | 0.90 (0.76-1.06) | 0.82 (0.65-1.04) | 1.02 (0.75-1.40) |
| DBP | 1.11 (0.91-1.35) | 1.38 (0.95-2.02) | 1.08 (0.78-1.49) | 1.08 (0.83-1.42) | 1.09 (0.77-1.53) | 0.89 (0.60-1.34) |
| BBzP | 1.09 (0.95-1.24) | 1.15 (0.93-1.43) | 1.17 (0.94-1.46) | 0.92 (0.79-1.07) | 0.82 (0.67-1.00) | 1.07 (0.85-1.33) |
| DEHP | 1.09 (0.91-1.30) | 1.21 (0.90-1.62) | 1.19 (0.89-1.60) | 1.15 (0.90-1.48) | 1.13 (0.82-1.56) | 1.04 (0.66-1.64) |
| MCPP | 1.07 (0.99-1.15) | 1.09 (0.96-1.24) | 1.06 (0.92-1.23) | 1.11 (0.87-1.41) | 0.94 (0.63-1.39) | 1.21 (0.85-1.74) |
| BPA ^a^ | 0.78 (0.48-1.28) | 0.76 (0.31-1.85) | 1.15 (0.54-2.43) | – | – | – |
| BPS ^a^ | 0.95 (0.48-1.90) | 1.26 (0.45-3.55) | 1.32 (0.44-3.98) | – | – | – |
| BPF ^a^ | 1.48 (0.84-2.59) | 1.86 (0.73-4.71) | 1.08 (0.41-2.85) | – | – | – |
| QG-comp ^b^ | 1.18 (0.88-1.58) | 1.48 (0.80-2.75) | 1.14 (0.60-2.14) | 0.96 (0.71-1.30) | 0.77 (0.49-1.22) | 1.13 (0.75-1.70) |

** p < 0.05 for interaction.* ^†^ *0.05 ≤ p < 0.10 for interaction.*

*^a^ BPA in top quartile; BPS and BPF in top decile. ^b^ Quantile g-computation estimates reflect the effect of a simultaneous one-quartile increase in DEP, DBP, BBzP, DEHP, and MCPP.*

*Abbreviations: BBzP = butyl benzyl phthalate; BIS: Barwon Infant study; BPA: bisphenol A; BPF: bisphenol F; BPS: bisphenol S; CHILD: Canadian Healthy Infant Longitudinal Development study; DBP: dibutyl phthalate; DEP: diethyl phthalate; DEHP = di(2-ethylhexyl) phthalate; DMP: dimethyl phthalate; MCPP = mono(3-carboxypropyl) phthalate; QG-comp: quantile G-computation.*

### Table S8. Fully adjusted risk ratios (and 95% confidence intervals) for outcomes associated with phthalates and bisphenols

|  | **BIS (Prenatal exposure) ^a^** | | | **CHILD (Postnatal exposure) ^b^** | | |
| --- | --- | --- | --- | --- | --- | --- |
| **Asthma** | **Overall**  **(N = 761)** | **Non-atopic**  **(N = 333)** | **Atopic**  **(N = 133)** | **Overall**  **(N = 976)** | **Non-atopic**  **(N = 814)** | **Atopic**  **(N = 123)** |
| DMP | 0.99 (0.86-1.14) | 1.09 (0.84-1.43) | 0.94 (0.77-1.14) | – | – | – |
| DEP | 1.00 (0.91-1.11) | 1.02 (0.87-1.20) | 0.99 (0.84-1.16) | 1.13 (0.98-1.31) | 1.13 (0.97-1.32) | 1.30 (0.81-2.10) |
| DBP | 1.02 (0.86-1.21) | 1.13 (0.83-1.55) | 0.94 (0.70-1.26) | 1.40 (1.11-1.77) | 1.49 (1.14-1.96) | 1.05 (0.69-1.62) |
| BBzP | 1.00 (0.91-1.11) | 1.17 (0.97-1.41) | 1.07 (0.88-1.31) | 1.19 (0.99-1.44) | 1.23 (0.99-1.52) | 1.19 (0.83-1.72) |
| DEHP | 0.93 (0.78-1.12) | 1.05 (0.77-1.43) | 0.89 (0.66-1.18) | 1.28 (0.99-1.67) | 1.30 (0.94-1.80) | 1.19 (0.73-1.94) |
| MCPP | 0.98 (0.92-1.05) | 1.13 (0.99-1.29) * | 0.93 (0.84-1.04) * | 1.32 (1.05-1.66) | 1.31 (0.98-1.75) | 1.21 (0.80-1.85) |
| BPA ^c^ | 0.82 (0.54-1.26) | 1.19 (0.60-2.33) | 0.94 (0.44-1.98) | – | – | – |
| BPS ^c^ | 0.76 (0.40-1.47) | 0.34 (0.08-1.41) ^†^ | 1.93 (0.67-5.61) ^†^ | – | – | – |
| BPF ^c^ | 1.34 (0.79-2.25) | 1.14 (0.45-2.88) | 1.95 (0.88-4.31) | – | – | – |
| QG-comp ^d^ | 0.99 (0.78-1.26) | 1.84 (1.11-3.04) | 0.79 (0.44-1.40) | 1.68 (1.20-2.36) | 1.81 (1.19-2.73) | 1.36 (0.74-2.48) |
| **Wheeze** | **Overall**  **(N = 767)** | **Non-atopic**  **(N = 331)** | **Atopic**  **(N = 134)** | **Overall**  **(N = 857)** | **Non-atopic**  **(N = 697)** | **Atopic**  **(N = 110)** |
| DMP | 0.98 (0.87-1.11) | 1.04 (0.82-1.32) | 0.97 (0.81-1.17) | – | – | – |
| DEP | 0.98 (0.90-1.06) | 1.02 (0.89-1.16) | 0.90 (0.79-1.03) | 1.20 (1.07-1.35) | 1.21 (1.07-1.38) | 1.19 (0.74-1.92) |
| DBP | 0.94 (0.82-1.08) | 0.96 (0.75-1.23) | 1.03 (0.79-1.34) | 1.40 (1.11-1.76) | 1.52 (1.16-2.00) | 1.03 (0.67-1.57) |
| BBzP | 0.98 (0.91-1.07) | 1.03 (0.88-1.21) | 1.07 (0.91-1.26) | 1.08 (0.92-1.26) | 1.06 (0.88-1.26) | 1.28 (0.88-1.85) |
| DEHP | 1.00 (0.87-1.14) | 1.00 (0.80-1.24) | 1.13 (0.84-1.50) | 1.09 (0.84-1.40) | 1.15 (0.85-1.56) | 0.95 (0.55-1.65) |
| MCPP | 0.99 (0.94-1.05) | 0.97 (0.88-1.07) | 1.04 (0.93-1.16) | 1.18 (0.94-1.48) | 1.31 (1.03-1.67) | 0.87 (0.55-1.38) |
| BPA ^c^ | 0.64 (0.44-0.93) | 0.65 (0.34-1.25) | 0.85 (0.42-1.72) | – | – | – |
| BPS ^c^ | 0.87 (0.52-1.45) | 0.54 (0.19-1.57) | 1.10 (0.34-3.57) | – | – | – |
| BPF ^c^ | 0.88 (0.55-1.41) | 0.70 (0.29-1.68) ^†^ | 1.80 (0.92-3.51) ^†^ | – | – | – |
| QG-comp ^d^ | 0.95 (0.78-1.15) | 1.07 (0.69-1.66) | 1.01 (0.57-1.80) | 1.53 (1.11-2.10) | 1.67 (1.16-2.39) | 1.16 (0.56-2.38) |
| **Eczema** | **Overall**  **(N = 729)** | **Non-atopic**  **(N = 298)** | **Atopic**  **(N = 122)** | **Overall**  **(N = 857)** | **Non-atopic**  **(N = 698)** | **Atopic**  **(N = 109)** |
| DMP | 1.05 (0.88-1.26) | 1.18 (0.92-1.50) | 1.03 (0.76-1.38) | – | – | – |
| DEP | 1.03 (0.93-1.14) | 1.03 (0.87-1.23) | 1.08 (0.90-1.29) | 0.96 (0.85-1.09) | 0.97 (0.84-1.13) | 0.92 (0.65-1.30) |
| DBP | 0.94 (0.79-1.12) | 0.92 (0.68-1.25) | 0.93 (0.68-1.28) | 0.96 (0.79-1.18) | 0.94 (0.74-1.20) | 0.81 (0.58-1.14) |
| BBzP | 1.01 (0.90-1.15) | 1.01 (0.81-1.25) | 1.11 (0.87-1.40) | 1.15 (1.03-1.28) | 1.12 (0.99-1.28) | 1.18 (0.93-1.50) |
| DEHP | 0.90 (0.72-1.12) | 0.90 (0.61-1.33) | 1.06 (0.72-1.54) | 0.97 (0.79-1.19) | 0.98 (0.78-1.23) | 0.83 (0.47-1.47) |
| MCPP | 1.06 (0.98-1.14) | 1.13 (0.98-1.29) | 0.99 (0.86-1.15) | 1.04 (0.89-1.23) | 1.06 (0.88-1.29) | 1.05 (0.71-1.55) |
| BPA ^c^ | 0.67 (0.40-1.12) | 0.80 (0.37-1.75) | 0.50 (0.13-1.92) | – | – | – |
| BPS ^c^ | 0.88 (0.45-1.70) | 1.71 (0.75-3.90) | 0.35 (0.03-3.48) | – | – | – |
| BPF ^c^ | 0.40 (0.15-1.04) | 0.23 (0.03-1.77) | 0.48 (0.12-1.96) | – | – | – |
| QG-comp ^d^ | 1.00 (0.76-1.30) | 1.09 (0.66-1.78) | 1.01 (0.50-2.00) | 1.05 (0.84-1.31) | 0.98 (0.77-1.25) | 1.09 (0.68-1.75) |
| **Rhinitis** | **Overall**  **(N = 695)** | **Non-atopic**  **(N = 330)** | **Atopic**  **(N = 133)** | **Overall**  **(N = 976)** | **Non-atopic**  **(N = 814)** | **Atopic**  **(N = 123)** |
| DMP | 1.07 (0.90-1.28) | 1.04 (0.85-1.28) | 1.09 (0.77-1.53) | – | – | – |
| DEP | 0.98 (0.87-1.10) | 0.97 (0.80-1.18) | 0.88 (0.73-1.06) | 0.91 (0.76-1.10) | 0.83 (0.66-1.06) | 1.11 (0.74-1.66) |
| DBP | 1.04 (0.85-1.29) | 1.27 (0.87-1.84) | 1.09 (0.70-1.70) | 1.08 (0.82-1.43) | 1.09 (0.80-1.49) | 0.94 (0.60-1.49) |
| BBzP | 1.09 (0.95-1.25) | 1.18 (0.95-1.46) | 1.21 (0.93-1.57) | 1.01 (0.84-1.22) | 0.89 (0.71-1.12) ^†^ | 1.24 (0.93-1.64) ^†^ |
| DEHP | 1.07 (0.88-1.31) | 1.15 (0.92-1.43) | 1.26 (0.88-1.80) | 1.13 (0.86-1.49) | 1.11 (0.78-1.57) | 1.03 (0.62-1.71) |
| MCPP | 1.05 (0.96-1.14) | 1.11 (0.96-1.28) | 1.07 (0.90-1.26) | 1.16 (0.90-1.50) | 0.96 (0.65-1.43) | 1.25 (0.81-1.92) |
| BPA ^c^ | 0.72 (0.43-1.22) | 0.72 (0.29-1.80) | 1.08 (0.42-2.79) | – | – | – |
| BPS ^c^ | 0.96 (0.46-2.00) | 1.33 (0.46-3.88) | 1.28 (0.38-4.33) | – | – | – |
| BPF ^c^ | 1.45 (0.78-2.68) | 1.71 (0.64-4.51) | 1.09 (0.32-3.73) | – | – | – |
| QG-comp ^d^ | 1.11 (0.83-1.50) | 1.40 (0.75-2.63) | 1.09 (0.52-2.32) | 1.04 (0.75-1.45) | 0.83 (0.51-1.35) | 1.24 (0.77-2.01) |

** p < 0.05 for interaction.* ^†^ *0.05 ≤ p < 0.10 for interaction.*

*^a^ Adjusted for* *age, sex, ethnicity, year and season of birth, family history of asthma, household income, SEIFA, household size, western diet during pregnancy, volatile household product score, and tobacco smoke exposure during pregnancy, pet ownership during pregnancy, living in a major Australian city, and house renovations during pregnancy.* ^b^ *Adjusted for centre of recruitment, age, sex, ethnicity, year and season of birth, family history of asthma, pet ownership at age five, household income, household size, breastfeeding duration, tobacco smoke exposure during pregnancy, house renovations at age five, and birthweight. ^c^ BPA in top quartile; BPS and BPF in top decile. ^d^ Quantile g-computation estimates reflect the effect of a simultaneous one-quartile increase in DEP, DBP, BBzP, DEHP, and MCPP.*

### Table S9. Risk ratios (and 95% confidence intervals) for challenge-proven food allergy associated with prenatal phthalates and bisphenols in BIS (N = 689)

|  | Crude | Minimally adjusted ^a^ | Fully adjusted ^b^ |
| --- | --- | --- | --- |
| DMP | 0.85 (0.72-1.00) | 0.85 (0.71-1.03) | 0.87 (0.71-1.06) |
| DEP | 0.99 (0.84-1.17) | 1.00 (0.84-1.20) | 1.03 (0.85-1.24) |
| DBP | 0.95 (0.75-1.21) | 0.94 (0.75-1.19) | 0.95 (0.75-1.20) |
| BBzP | 0.89 (0.74-1.07) | 0.88 (0.73-1.05) | 0.88 (0.73-1.06) |
| DEHP | 0.84 (0.63-1.11) | 0.84 (0.63-1.12) | 0.84 (0.62-1.14) |
| MCPP | 0.96 (0.87-1.05) | 0.97 (0.87-1.07) | 0.97 (0.87-1.09) |
| BPA ^c^ | 0.12 (0.03-0.47) | 0.12 (0.03-0.49) | 0.13 (0.03-0.53) |
| BPS ^c^ | 0.34 (0.08-1.40) | 0.32 (0.07-1.42) | 0.32 (0.07-1.43) |
| BPF ^c^ | 1.28 (0.60-2.76) | 1.36 (0.61-3.06) | 1.49 (0.65-3.44) |
| QG-comp ^d^ | 0.79 (0.55-1.15) | 0.79 (0.55-1.13) | 0.82 (0.55-1.21) |

.*^a^ Adjusted for age, sex, ethnicity, year and season of birth, family history of asthma, household income, SEIFA, household size, western diet during pregnancy, volatile household product score, and tobacco smoke exposure during pregnancy.*

*^b^ Model further adjusted for pet ownership during pregnancy, living in a major Australian city, and house renovations during pregnancy.*

*^c^ BPA in top quartile; BPS and BPF in top decile.*

*^d^ Quantile g-computation estimates reflect the effect of a simultaneous one-quartile increase in DEP, DBP, BBzP, DEHP, and MCPP.*

*Abbreviations: BBzP = butyl benzyl phthalate; BIS: Barwon Infant study; BPA: bisphenol A; BPF: bisphenol F; BPS: bisphenol S; CHILD: Canadian Healthy Infant Longitudinal Development study; DBP: dibutyl phthalate; DEP: diethyl phthalate; DEHP = di(2-ethylhexyl) phthalate; DMP: dimethyl phthalate; MCPP = mono(3-carboxypropyl) phthalate; QG-comp: quantile G-computation.*

### Table S10. Adjusted risk ratios (and 95% confidence intervals) for HDM/grass sensitisation associated with prenatal phthalates and bisphenols in BIS

|  | **BIS (Prenatal exposure; N = 465) ^a^** | | | **CHILD (Postnatal exposure; N = 930) ^b^** | | |
| --- | --- | --- | --- | --- | --- | --- |
|  | **Grass exclusive**  **(Cases = 29)** | **HDM**  **exclusive**  **(Cases = 54)** | **HDM and grass sensitisation**  **(Cases = 37)** | **Grass exclusive**  **(Cases = 26)** | **HDM**  **exclusive**  **(Cases = 27)** | **HDM and grass sensitisation**  **(Cases = 13)** |
| DMP | 0.76 (0.56-1.03) | 1.14 (0.93-1.39) | 0.69 (0.52-0.92) | – | – | – |
| DEP | 1.07 (0.88-1.31) | 0.96 (0.83-1.10) | 0.97 (0.80-1.18) | 1.00 (0.74-1.36) | 0.92 (0.70-1.19) | 0.94 (0.48-1.83) |
| DBP | 0.86 (0.53-1.38) | 0.98 (0.72-1.33) | 1.17 (0.83-1.64) | 0.96 (0.54-1.69) | 1.17 (0.68-2.03) | 0.80 (0.33-1.96) |
| BBzP | 0.75 (0.57-0.98) | 0.91 (0.75-1.10) | 0.92 (0.70-1.20) | 1.03 (0.80-1.33) | 1.29 (0.96-1.72) | 0.66 (0.41-1.08) |
| DEHP | 0.75 (0.46-1.23) | 1.00 (0.78-1.28) | 0.97 (0.74-1.25) | 0.92 (0.58-1.47) | 1.28 (0.80-2.06) | 1.45 (0.63-3.33) |
| MCPP | 0.96 (0.83-1.10) | 1.00 (0.89-1.11) | 0.98 (0.87-1.11) | 0.95 (0.62-1.44) | 1.13 (0.78-1.63) | 2.31 (1.11-4.85) |
| BPA ^c^ | 0.78 (0.32-1.90) | 0.96 (0.54-1.71) | 0.41 (0.14-1.20) | – | – | – |
| BPS ^c^ | 0.74 (0.20-2.70) | 0.87 (0.39-1.94) | NC ^e^ | – | – | – |
| BPF ^c^ | 1.11 (0.26-4.79) | 0.98 (0.40-2.39) | 0.69 (0.17-2.81) | – | – | – |
| QG-comp ^d^ | 0.71 (0.42-1.19) | 0.98 (0.67-1.43) | 1.00 (0.64-1.56) | 0.96 (0.55-1.69) | 1.39 (0.80-2.41) | 0.89 (0.38-2.06) |

*^a^ Adjusted for age, sex, ethnicity, year and season of birth, family history of asthma, household income, SEIFA, household size, western diet during pregnancy, volatile household product score, tobacco smoke exposure during pregnancy, and season of skin prick testing.*

^b^ *Adjusted for centre of recruitment, age, sex, ethnicity, year and season of birth, family history of asthma, pet ownership at age five, household income, household size, breastfeeding duration, tobacco smoke exposure during pregnancy, and season of skin prick testing.*

*^c^ BPA in top quartile; BPS and BPF in top decile.*

*^d^ Quantile g-computation estimates reflect the effect of a simultaneous one-quartile increase in DEP, DBP, BBzP, DEHP, and MCPP.*

*^e^ Non-convergence due to absence of exposed cases*

*Abbreviations: BBzP = butylbenzyl phthalate; BIS: Barwon Infant study; BPA: bisphenol A; BPF: bisphenol F; BPS: bisphenol S; CHILD: Canadian Healthy Infant Longitudinal Development study; DBP: dibutyl phthalate; DEP: diethyl phthalate; DEHP = di(2-ethylhexyl) phthalate; DMP: dimethyl phthalate; MCPP = mono(3-carboxypropyl) phthalate; QG-comp: quantile G-computation.*

### Table S11. Adjusted difference in means (and 95% confidence intervals) for lung function outcomes associated with prenatal phthalates and bisphenols in BIS

|  | **BIS (Prenatal exposure) ^a^** | | | **CHILD (Postnatal exposure) ^b^** | | |
| --- | --- | --- | --- | --- | --- | --- |
| **FRC** | **Overall**  **(N = 205)** | **Non-atopic**  **(N = 126)** | **Atopic**  **(N = 45)** | **Overall**  **(N = 54)** | **Non-atopic**  **(N = 43)** | **Atopic**  **(N = 5)** |
| DMP | 0.01 (-0.08, 0.10) | -0.02 (-0.16, 0.13) | 0.01 (-0.10, 0.11) | – | – | – |
| DEP | 0.01 (-0.06, 0.07) | -0.02 (-0.13, 0.09) | 0.05 (-0.06, 0.16) | -0.09 (-0.33, 0.14) | 0.03 (-0.32, 0.38) | -0.64 (-1.75, 0.47) |
| DBP | -0.01 (-0.14, 0.11) | -0.12 (-0.31, 0.06) | 0.00 (-0.26, 0.27) | -0.03 (-0.33, 0.26) | -0.16 (-0.56, 0.23) | 0.13 (-1.95, 2.21) |
| BBzP | 0.01 (-0.07, 0.09) | -0.08 (-0.18, 0.03) | 0.02 (-0.12, 0.16) | -0.13 (-0.42, 0.17) | -0.23 (-0.65, 0.19) | -0.45 (-1.56, 0.67) |
| DEHP | 0.03 (-0.11, 0.16) | 0.00 (-0.21, 0.21) | 0.02 (-0.27, 0.30) | 0.38 (0.10, 0.67) | 0.34 (-0.18, 0.86) | 0.03 (-0.88, 0.94) |
| MCPP | -0.02 (-0.07, 0.03) | -0.04 (-0.12, 0.03) | 0.06 (-0.03, 0.15) | 0.29 (0.03, 0.55) | 0.22 (-0.42, 0.86) | 0.20 (-1.68, 2.09) |
| BPA ^c^ | 0.07 (-0.21, 0.35) | -0.08 (-0.43, 0.28) | 0.54 (-0.12, 1.19) | – | – | – |
| BPS ^c^ | 0.06 (-0.36, 0.47) | 0.06 (-0.50, 0.61) | 0.16 (-1.30, 1.62) | – | – | – |
| BPF ^c^ | 0.15 (-0.24, 0.54) | 0.05 (-0.49, 0.59) | 0.25 (-0.47, 0.97) | – | – | – |
| QG-comp ^d^ | -0.03 (-0.18, 0.13) | -0.18 (-0.37, 0.02) * | 0.27 (0.02, 0.53) * | NC ^e^ | NC ^e^ | NC ^e^ |
| **LCI_2.5_** | **Overall**  **(N = 205)** | **Non-atopic**  **(N = 126)** | **Atopic**  **(N = 45)** | **Overall**  **(N = 54)** | **Non-atopic**  **(N = 43)** | **Atopic**  **(N = 5)** |
| DMP | 0.02 (-0.09, 0.12) | 0.06 (-0.17, 0.29) | 0.02 (-0.15, 0.19) | – | – | – |
| DEP | -0.01 (-0.09, 0.07) | 0.00 (-0.13, 0.12) | -0.07 (-0.21, 0.07) | 0.03 (-0.20, 0.26) | 0.01 (-0.33, 0.36) | 0.34 (-1.18, 1.87) |
| DBP | 0.05 (-0.10, 0.20) | 0.15 (-0.11, 0.41) | 0.00 (-0.31, 0.32) | 0.05 (-0.25, 0.35) | 0.08 (-0.38, 0.53) | 0.37 (-1.43, 2.16) |
| BBzP | 0.00 (-0.09, 0.09) | 0.04 (-0.11, 0.19) | 0.02 (-0.17, 0.21) | 0.23 (-0.05, 0.51) | 0.28 (-0.13, 0.70) | 0.58 (-1.80, 2.95) |
| DEHP | -0.02 (-0.18, 0.14) | 0.00 (-0.23, 0.22) | 0.05 (-0.32, 0.42) | -0.21 (-0.51, 0.09) | -0.27 (-0.77, 0.23) | 0.33 (-0.59, 1.25) |
| MCPP | -0.01 (-0.07, 0.04) | 0.00 (-0.08, 0.08) | -0.05 (-0.16, 0.06) | -0.07 (-0.35, 0.21) | 0.09 (-0.70, 0.87) | 0.15 (-1.68, 1.98) |
| BPA ^c^ | 0.02 (-0.32, 0.35) | -0.04 (-0.46, 0.37) | -0.07 (-1.09, 0.94) | – | – | – |
| BPS ^c^ | -0.12 (-0.61, 0.37) | 0.01 (-0.82, 0.83) | -0.72 (-2.93, 1.49) | – | – | – |
| BPF ^c^ | 0.33 (-0.14, 0.79) | 0.30 (-0.43, 1.03) | 0.42 (-0.73, 1.56) | – | – | – |
| QG-comp ^d^ | -0.03 (-0.24, 0.17) | 0.06 (-0.19, 0.32) | -0.19 (-0.56, 0.18) | NC ^e^ | NC ^e^ | NC ^e^ |
| **Resistance** | **Overall**  **(N = 393)** | **Non-atopic**  **(N = 214)** | **Atopic**  **(N = 45)** | **Overall**  **(N = 154)** | **Non-atopic**  **(N = 111)** | **Atopic**  **(N = 23)** |
| DMP | -0.02 (-0.10, 0.06) | 0.00 (-0.10, 0.10) | -0.05 (-0.16, 0.07) | – | – | – |
| DEP | -0.04 (-0.09, 0.01) | 0.03 (-0.05, 0.12) * | -0.20 (-0.41, 0.00) * | 0.01 (-0.10, 0.12) | 0.10 (-0.03, 0.23) | 0.05 (-0.13, 0.24) |
| DBP | -0.03 (-0.14, 0.07) | 0.02 (-0.11, 0.15) | -0.10 (-0.31, 0.12) | 0.11 (-0.06, 0.28) | 0.10 (-0.11, 0.31) | 0.17 (-0.21, 0.55) |
| BBzP | -0.05 (-0.11, 0.02) | -0.02 (-0.11, 0.07) | -0.04 (-0.16, 0.08) | 0.02 (-0.12, 0.16) | -0.03 (-0.18, 0.11) | 0.23 (-0.17, 0.62) |
| DEHP | -0.02 (-0.12, 0.09) | 0.02 (-0.09, 0.13) | -0.09 (-0.26, 0.08) | 0.10 (-0.09, 0.29) | 0.10 (-0.18, 0.38) | 0.28 (-0.06, 0.63) |
| MCPP | 0.01 (-0.03, 0.04) | -0.02 (-0.07, 0.03) | 0.00 (-0.08, 0.07) | 0.09 (-0.08, 0.27) | 0.04 (-0.17, 0.25) ^†^ | 0.46 (0.01, 0.91) ^†^ |
| BPA ^c^ | -0.21 (-0.43, 0.01) | -0.30 (-0.57, -0.03) | -0.18 (-0.69, 0.33) | – | – | – |
| BPS ^c^ | 0.09 (-0.25, 0.43) | 0.10 (-0.35, 0.55) | 0.29 (-0.41, 0.98) | – | – | – |
| BPF ^c^ | -0.13 (-0.44, 0.18) | -0.34 (-0.81, 0.14) * | 0.43 (-0.15, 1.00) * | – | – | – |
| QG-comp ^d^ | -0.09 (-0.22, 0.04) | 0.04 (-0.14, 0.22) * | -0.34 (-0.58, -0.09) * | 0.11 (-0.09, 0.30) | 0.16 (-0.08, 0.41) | 0.41 (-0.03, 0.85) |
| **Reactance** | **Overall**  **(N = 393)** | **Non-atopic**  **(N = 214)** | **Atopic**  **(N = 45)** | **Overall**  **(N = 154)** | **Non-atopic**  **(N = 111)** | **Atopic**  **(N = 23)** |
| DMP | -0.03 (-0.10, 0.05) | 0.01 (-0.08, 0.11) | -0.07 (-0.20, 0.06) | – | – | – |
| DEP | 0.00 (-0.06, 0.05) | -0.04 (-0.13, 0.04) * | 0.09 (-0.03, 0.21) * | -0.02 (-0.14, 0.10) | -0.06 (-0.24, 0.11) | -0.14 (-0.34, 0.05) |
| DBP | 0.00 (-0.10, 0.11) | -0.03 (-0.15, 0.10) | 0.05 (-0.22, 0.32) | -0.18 (-0.36, 0.00) | -0.23 (-0.44, -0.03) | -0.05 (-0.53, 0.43) |
| BBzP | -0.01 (-0.07, 0.06) | -0.05 (-0.13, 0.03) | 0.02 (-0.10, 0.14) | -0.04 (-0.19, 0.11) | 0.02 (-0.12, 0.16) | -0.17 (-0.45, 0.12) |
| DEHP | -0.06 (-0.16, 0.05) | -0.09 (-0.21, 0.02) | -0.06 (-0.25, 0.13) | -0.08 (-0.29, 0.12) | -0.27 (-0.57, 0.02) | -0.06 (-0.38, 0.25) |
| MCPP | 0.00 (-0.04, 0.04) | 0.00 (-0.04, 0.04) | 0.02 (-0.06, 0.10) | -0.06 (-0.25, 0.13) | -0.08 (-0.27, 0.10) | -0.18 (-0.69, 0.32) |
| BPA ^c^ | 0.07 (-0.15, 0.30) | 0.29 (0.06, 0.52) | -0.12 (-0.77, 0.53) | – | – | – |
| BPS ^c^ | -0.20 (-0.54, 0.14) | 0.08 (-0.45, 0.60) | -0.66 (-1.55, 0.22) | – | – | – |
| BPF ^c^ | -0.01 (-0.33, 0.30) | 0.12 (-0.26, 0.51) | -0.36 (-1.13, 0.40) | – | – | – |
| QG-comp ^d^ | -0.02 (-0.14, 0.10) | -0.13 (-0.28, 0.02) ^†^ | 0.19 (-0.11, 0.49) ^†^ | -0.16 (-0.36, 0.05) | -0.27 (-0.51, -0.03) | -0.32 (-0.71, 0.07) |

** p < 0.05 for interaction.* ^†^ *0.05 ≤ p < 0.10 for interaction.*

*^a^ Adjusted for age, sex, height, ethnicity, year and season of birth, family history of asthma, household income, SEIFA, household size, western diet during pregnancy, volatile household product score, and tobacco smoke exposure during pregnancy.*

^b^ *Adjusted for centre of recruitment, age, sex, height, ethnicity, year and season of birth, family history of asthma, pet ownership at age five, household income, household size, breastfeeding duration and tobacco smoke exposure during pregnancy.*

*^c^ BPA in top quartile; BPS and BPF in top decile.*

*^d^ Quantile g-computation estimates reflect the effect of a simultaneous one-quartile increase in DEP, DBP, BBzP, DEHP, and MCPP.*

*^e^ Non-convergence due to small sample size.*

*Abbreviations: BBzP = butyl benzyl phthalate; BIS: Barwon Infant study; BPA: bisphenol A; BPF: bisphenol F; BPS: bisphenol S; CHILD: Canadian Healthy Infant Longitudinal Development study; DBP: dibutyl phthalate; DEP: diethyl phthalate; DEHP = di(2-ethylhexyl) phthalate; DMP: dimethyl phthalate; MCPP = mono(3-carboxypropyl) phthalate; QG-comp: quantile G-computation.*

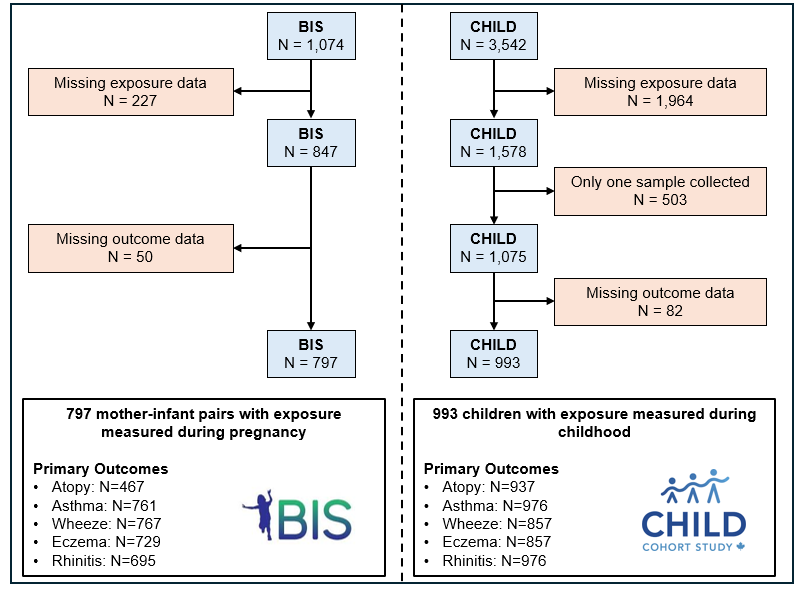

### Figure S1. Population inclusion flowchart

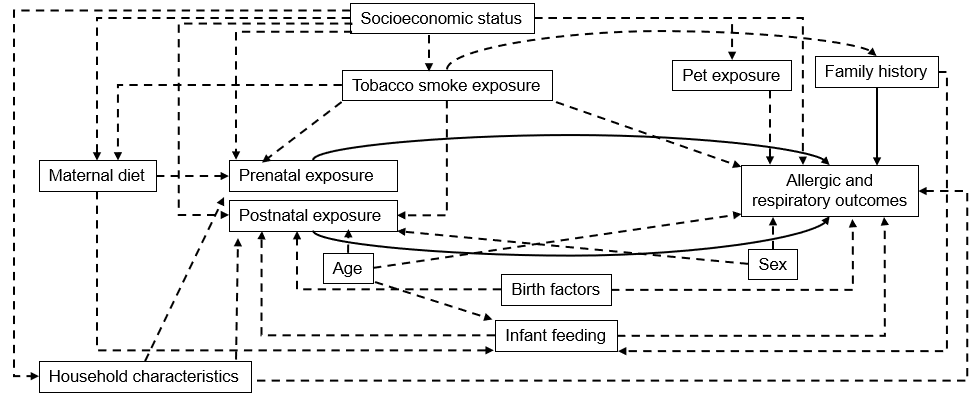

### Figure S2. Simplified Directed Acyclic Graph depicting the effect of phthalates and bisphenols on childhood allergies

*Solid arrows represent causal pathways.*

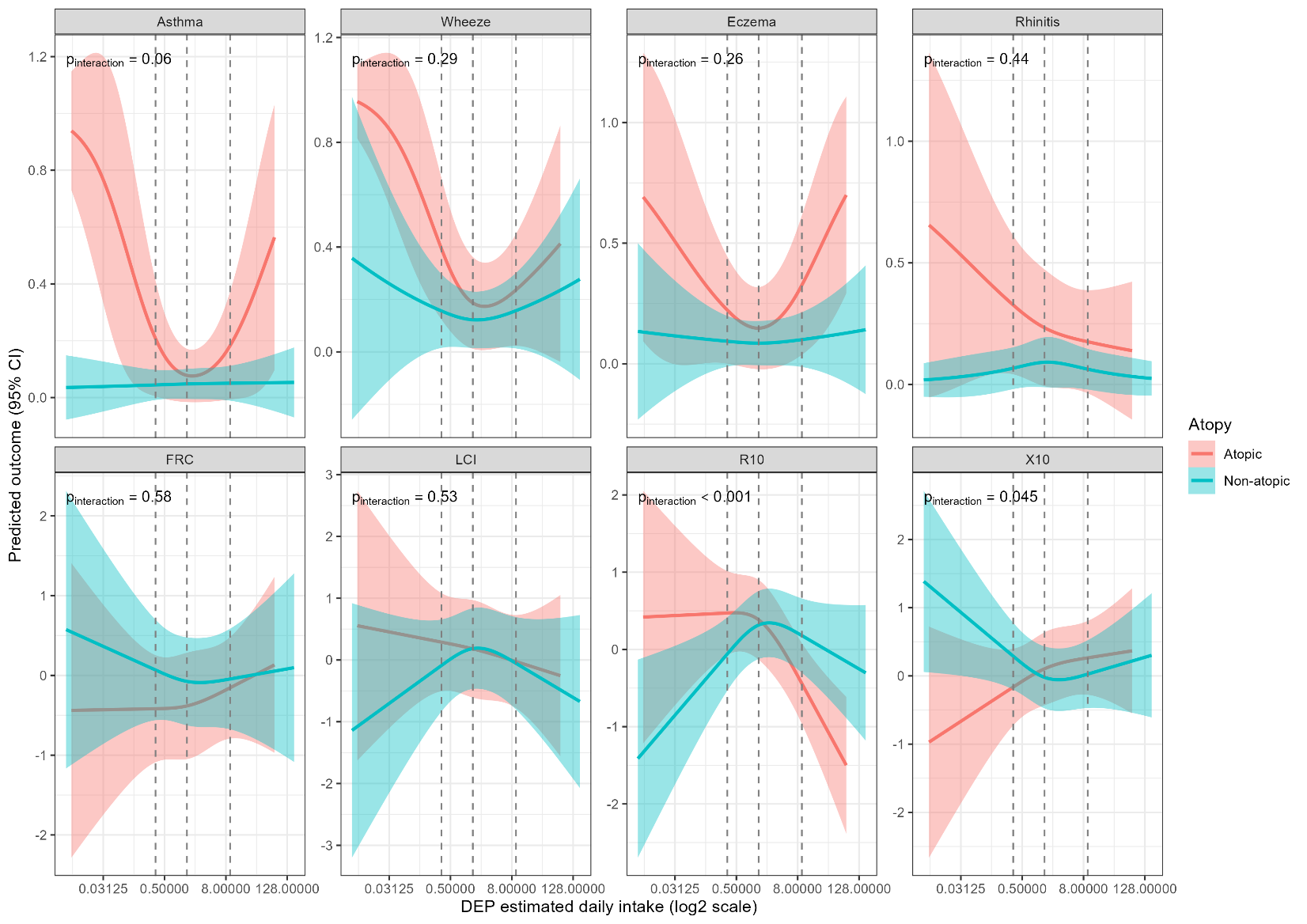

### Figure S3. Dose–response relationships between prenatal DEP exposure and allergic and lung function outcomes in BIS, modelled using restricted cubic splines

*The Y-axis represents scaled outcomes for continuous outcomes and probabilities for binary outcomes. Vertical dashed lines represent DEP’s 10, 50^th^ and 90^th^ percentiles. Adjusted for age, sex, ethnicity, year and season of birth, family history of asthma, household income, SEIFA, household size, western diet during pregnancy, volatile household product score, and tobacco smoke exposure during pregnancy. Lung function models were further adjusted for height.*

*Abbreviations: DEP: Di-ethyl phthalate; FRC: Function residual capacity; LCI: Lung clearance index; R10: Resistance at 10Hz; X10: Reactance at 10Hz.*

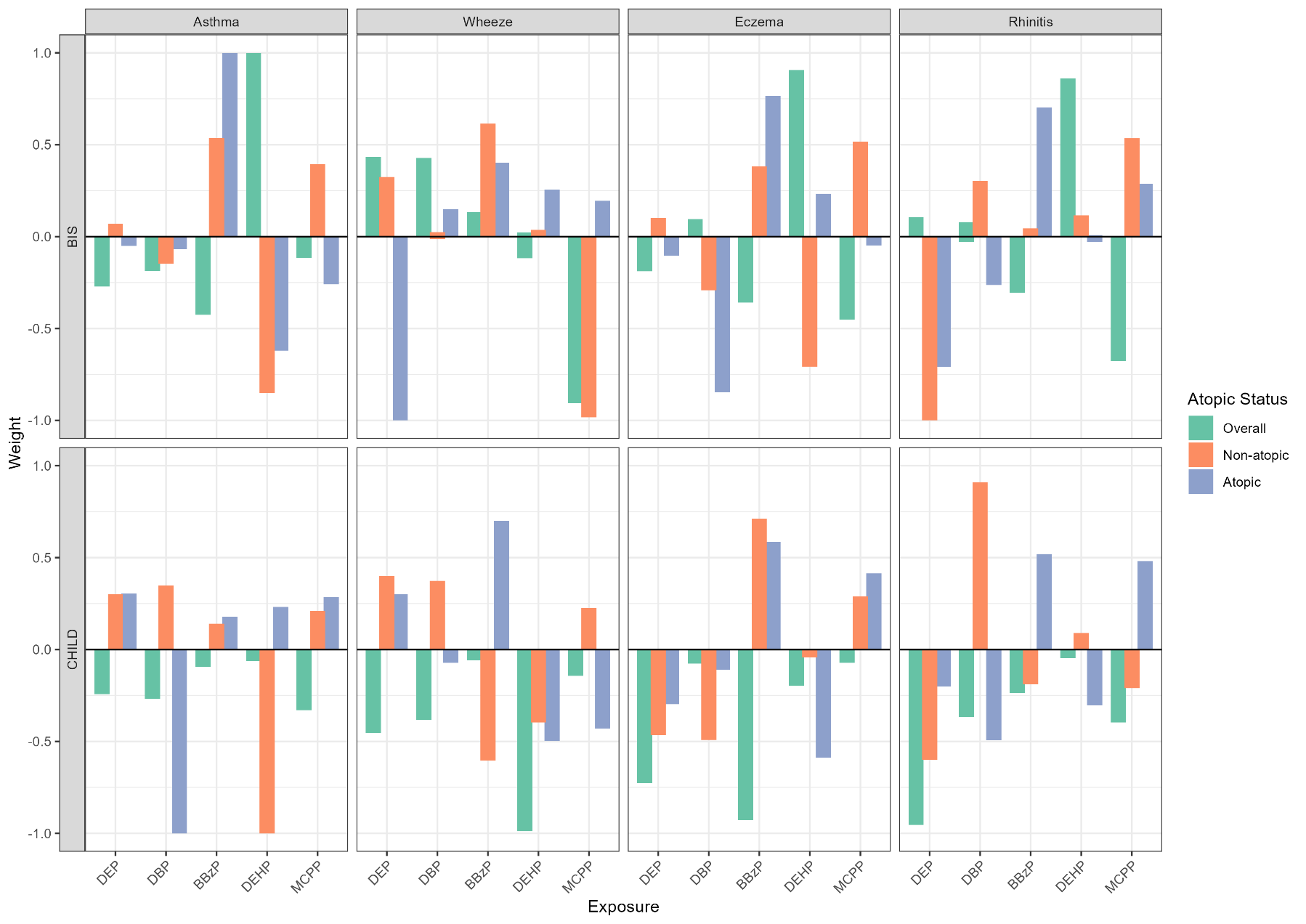

### Figure S4. Relative weight of individual phthalates in quantile g-computation models

*Within each direction weights are constrained to sum one.*

*Abbreviations: BBzP = butylbenzyl phthalate; BIS: Barwon Infant study; BPA: bisphenol A; BPF: bisphenol F; BPS: bisphenol S; CHILD: Canadian Healthy Infant Longitudinal Development study; DBP: dibutyl phthalate; DEP: diethyl phthalate; DEHP = di(2-ethylhexyl) phthalate; MCPP = mono(3-carboxypropyl) phthalate*

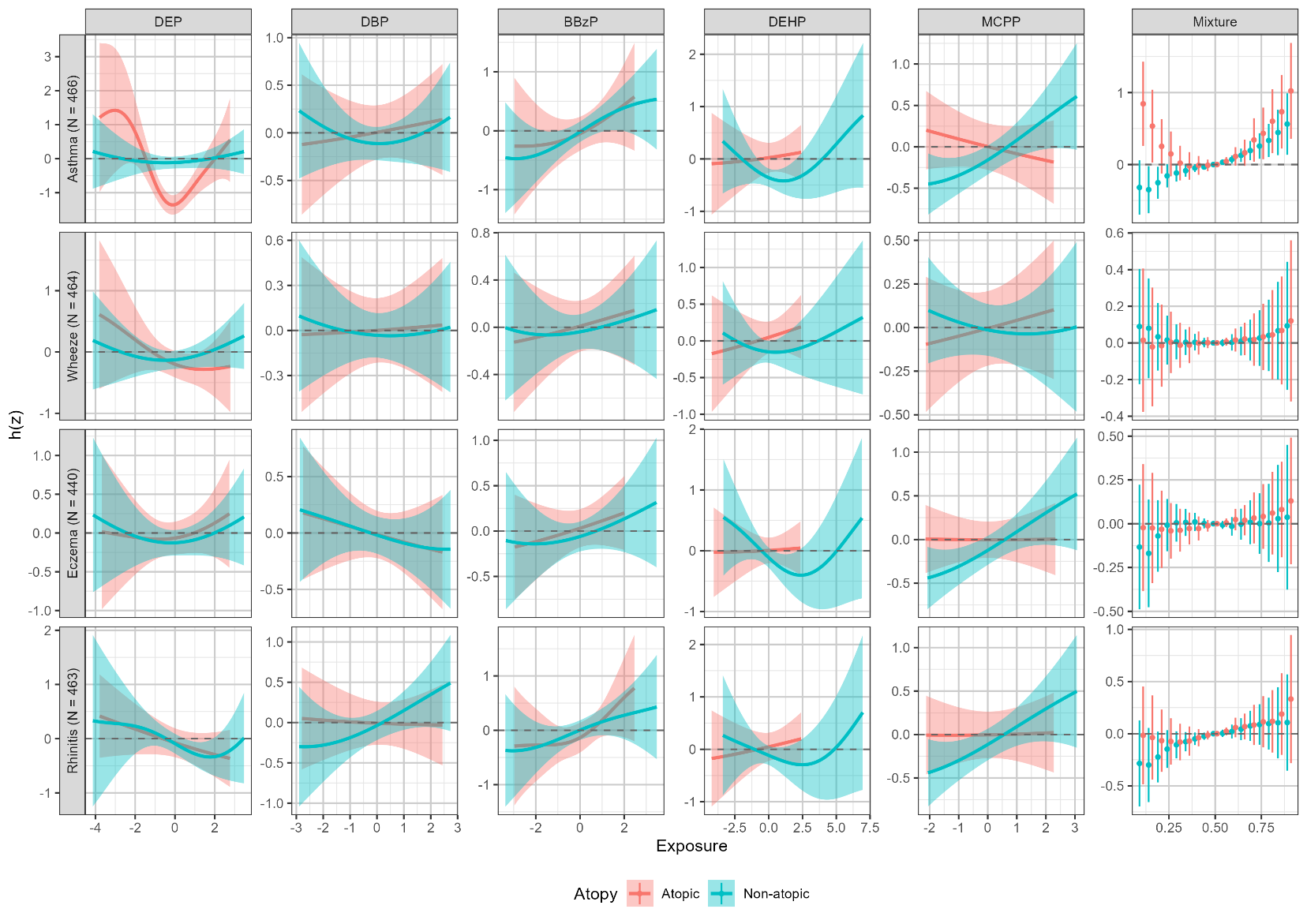

### Figure S5. Multivariate dose–response relationships between prenatal exposures and allergic outcomes in BIS, stratified by atopy, using BKMR

*The x-axis shows exposure z-score for single compounds, while it represents joint exposure quantiles for the mixture. Dose responses for individual compounds are shown fixing other compounds at their median values.*

*Adjusted for age, sex, ethnicity, year and season of birth, family history of asthma, household income, SEIFA, household size, western diet during pregnancy, volatile household product score, and tobacco smoke exposure during pregnancy.*

*Abbreviations: BBzP = butyl benzyl phthalate; BIS: Barwon Infant study; DBP: dibutyl phthalate; DEP: diethyl phthalate; DEHP = di(2-ethylhexyl) phthalate; MCPP: mono(3-carboxypropyl) phthalate*

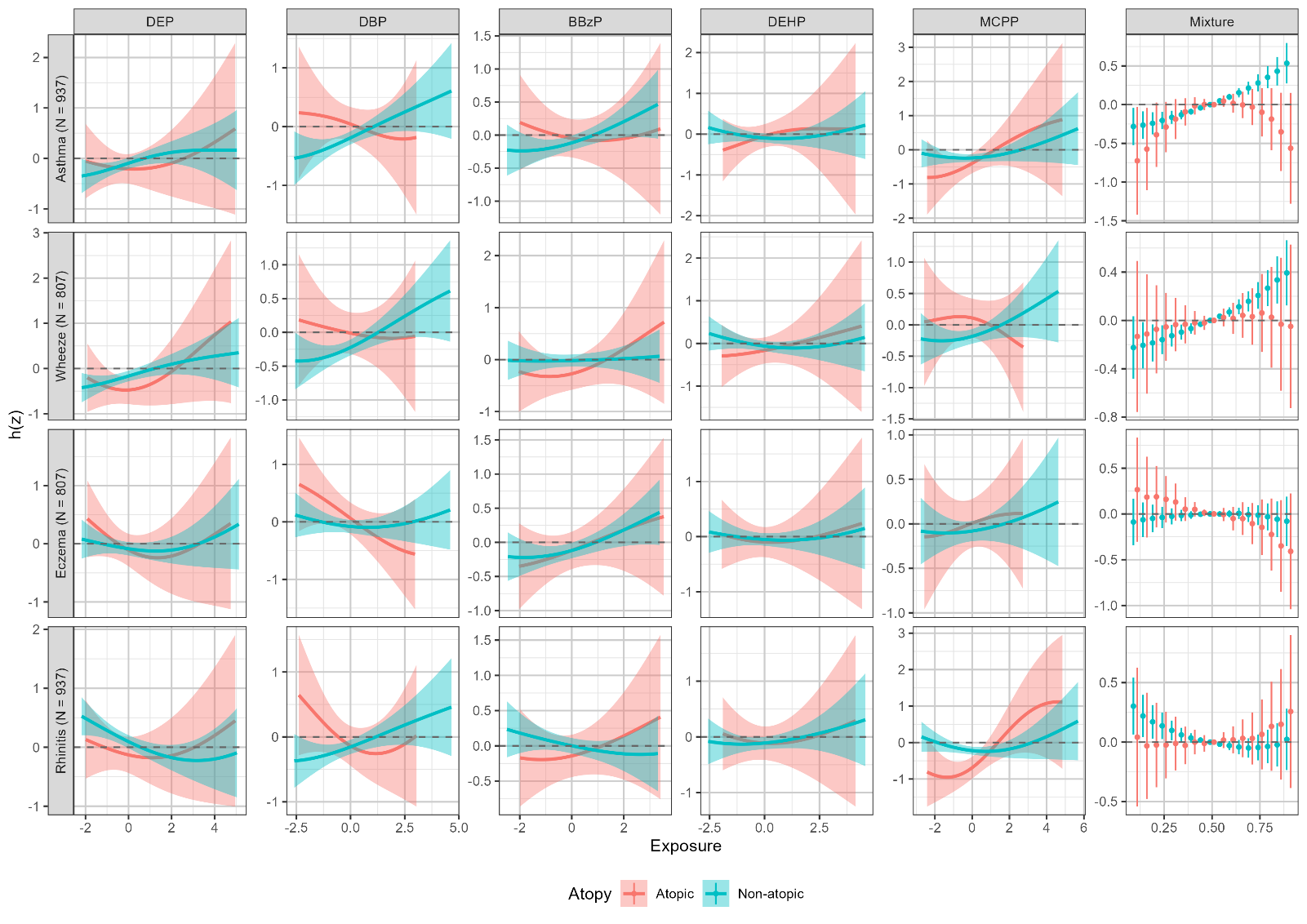

### Figure S6. Multivariate dose–response relationships between postnatal exposures and allergic outcomes in CHILD, stratified by atopy, using BKMR

*The x-axis shows exposure z-score for single compounds, while it represents joint exposure quantiles for the mixture. Dose responses for individual compounds are shown fixing other compounds at their median values*

*Adjusted for* *c**entre of recruitment, age, sex, ethnicity, year and season of birth, family history of asthma, pet ownership at age five, household income, household size, breastfeeding duration and tobacco smoke exposure during pregnancy.*

*Abbreviations: BBzP = butylbenzyl phthalate; BIS: Barwon Infant study; DBP: dibutyl phthalate; DEP: diethyl phthalate; DEHP = di(2-ethylhexyl) phthalate; MCPP: mono(3-carboxypropyl) phthalate*
